# Time for Tobacco Elimination: Modelling smoking cessation strategies and lung cancer screening in Singapore

**DOI:** 10.64898/2026.05.06.26352560

**Authors:** Yichen He, Shihui Jin, Xinyu Zhang, King Ian Fong, Yi Wang, Kelvin Bryan Tan, Ross Soo, Jue Tao Lim, Borame L Dickens

## Abstract

**Background:** Lung cancer remains a major public health burden, with poor survival largely driven by late-stage diagnosis. With declining and very low smoking prevalence in Singapore at 4.7% in 2024 among 18–29-year-olds, questions arise about future screening efficiency, eligibility criteria, and the impact of smoking cessation, including tobacco elimination.

**Methods:** We developed a large-scale microsimulation model calibrated to real-world data, generating individual life histories, smoking trajectories, and disease progression for Singapore’s 4.18 million residents to project smoking prevalence and lung cancer burden. We evaluated 271 low-dose computed tomography (LDCT) screening strategies (by age, gender, uptake, and frequency) under five tobacco control scenarios, from status quo to a complete smoking ban, between 2025 and 2050.

**Findings:** Under the status quo, all screening strategies were cost-effective relative to the 2024 GDP per capita threshold (∼SGD 120,000). Among strategies with ≤10% overdiagnosis, annual screening of eligible ever-smokers aged 50– 80 years was most life-saving, yielding 51,312 (95% uncertainty interval: 36,821–72,830) QALYs at a total cost of SGD 12.2 (9.7–16.1) billion. Adding an immediate smoking ban increased QALY gains by 2.8 (2.2–3.5) times while reducing the total cost by 23.3% (17.0%–30.0%). Extending eligibility to individuals with lower smoking exposure or a first-degree family history remained cost-effective.

**Interpretations:** Tobacco elimination yields substantial health and economic benefits, while well-designed risk-based LDCT screening of residual high-risk populations remains cost-effective, supporting a continued role for screening even in settings with declining smoking prevalence.

## 1. Introduction

Lung cancer constitutes a major global public health burden, with an estimated 2.5 million incident cases and approximately 1.8 million deaths annually worldwide.^1^ Prognosis remains poor with five-year survival rates typically ranging from 15% to 25%, largely reflecting late-stage diagnosis, which is a primary determinant of survival outcomes.^2–5^ High-risk populations are defined predominantly by tobacco smoking exposure, accounting for an estimated 80–85% of lung cancer deaths,^6,7^ yet a substantial proportion of lung cancer in Singapore occurs among never-smokers, particularly women, among whom approximately 60–80% of cases have no reported smoking history.^8^ Furthermore, national data indicate a five-year survival of approximately 21.9%, with late-stage disease accounting for around 80% of diagnoses in men and more than 70% in women, highlighting ongoing challenges for early detection via screening practices.^9^ Of particular concern is that a high proportion of lung cancer incidences, up to 61% and 83%, are reported among women in East and Southeast Asia, respectively.^10^

Currently, population-level screening strategies with low-dose CT (LDCT) have been recommended for eligible screening populations, as randomised controlled trials have demonstrated a 20% or greater reduction in lung cancer-specific mortality in these groups.^11^ Singapore’s screening recommendations follow the U.S. Preventive Services Task Force (USPSTF) to include adults aged 50 to 80 years with a smoking history of at least 20 pack years who currently smoke or who have quit within the past 15 years.^12,13^ This higher-risk population has been shrinking, however, with falling smoking prevalence from 18.3% in 1992 to 8.4% in 2024,^14^ reflecting sustained tobacco control efforts initiated with the National Smoking Control Programme in 1986. Efforts to reduce smoking prevalence in Singapore are continuing, with discussions around a generational tobacco ban aimed at creating a smoke-free generation. This follows Singapore’s long-standing ban on vaping, which has been in place since 2018 and was recently strengthened through the Tobacco and Vaporisers Control Act passed in March 2026,^15,16^ as well as a 20% increase in tobacco excise duty implemented on 12 February 2026.^17^ Given that Singapore has one of the lowest smoking prevalence in the world and relatively high institutional trust,^18,19^ it appears well placed to begin considering endgame measures such as the introduction of a smoking ban.

Several important uncertainties remain regarding the relative reduction in lung cancer burden achievable through national smoking cessation strategies compared with different lung cancer screening approaches, as well as the extent to which declining smoking prevalence will reshape the effectiveness and design of future screening programmes. The proportion of lung cancer cases occurring among individuals classified as never smokers is also expected to rise alongside population ageing.^1^ Furthermore, in a low smoking prevalence setting, it remains unclear whether individuals with lower cumulative smoking exposure, who nonetheless have an elevated risk relative to never smokers,^20^ should be included within screening eligibility thresholds as screening resources can be more widely distributed.

This study therefore aims to quantify the reduction in lung cancer burden under combined screening and smoking cessation scenarios, ranging from status quo to an immediate smoking ban, using a national microsimulation model from 2025 to 2050, considering screening programme costs, Quality-Adjusted Life Years (QALYs) gained, number of false positives, overdiagnoses, late-stage cases and lung cancer specific deaths averted. Factors such as screening age, gender, and different at-risk populations according to smoking behaviours and the presence of lung cancer first-degree family history are also explored. We (i) estimated the long-term impact of tobacco control relative to targeted screening in a low smoking prevalence context, (ii) evaluated how varying smoking cessation trajectories influenced the performance and cost effectiveness of screening strategies, and (iii) assessed the feasibility of extending risk-based screening to ever-smokers with limited smoking exposure and other at-risk populations.

## 2. Methods

### 2.1 Overview

We developed a large-scale microsimulation framework, calibrated and validated with empirical data on smoking behaviour and lung cancer epidemiology from national cohort studies and registries. The model generated individual smoking histories and disease progression for Singapore’s 4.18 million residents, projecting smoking prevalence, lung cancer incidence and mortality, and associated healthcare costs from 1990 to 2050.

We further evaluated the health and economic impacts of LDCT screening from 2025 to 2050 across 271 screening programme designs defined by age, gender, uptake, and screening frequency, under five tobacco control scenarios: status quo; mild, moderate, and stringent increases in smoking cessation relative to baseline (25%, 100%, and 400%, respectively); and a complete smoking ban from 2025. In the complete ban scenario, no new smoking initiation was assumed from 2025 onward and all current smokers were assumed to quit in 2025. Strategies were evaluated using projected screening-related benefits and harms to identify optimal policy combinations.

### 2.2 Synthetic Singapore Population

Individual life trajectories were derived from the Demographic Epidemiological Model of Singapore (DEMOS), a dynamic microsimulation framework that generates a virtual cohort of approximately 4.18 million Singapore residents (as of June 2024), characterised by detailed demographics, such as age, gender, ethnicity. The fertility and mortality rates in DEMOS were age-, gender-, and ethnicity-specific, projected based on the 2024 demographic statistics.^21^ We simulated each individual up to age 90 years. Further details on the DEMOS model are provided in Normatov et al.^22^

### 2.3 Smoking History Model

Individual smoking profiles over the lifespan were featured by both smoking states and intensity. The smoking states were updated annually from the earliest possible initiation age of 12 years. We considered three smoking states: never, current, and former smoker, where individuals may transition from never to current smoker, and subsequently quit, with transition probabilities determined by age, gender, ethnicity, and birth cohort. Relapse among former smokers was assumed not to occur. Each ever-smoker was assumed to maintain a constant smoking intensity throughout the smoking period. This smoking intensity, defined as the average number of cigarette packs smoked per day,^23^ was independently sampled from gender-specific empirical distributions derived from Singapore multi-ethnic cohort data (Figure S2).^24^

### 2.4 Natural History Model

We adapted the MIcroSimulation Cost-effectiveness ANalysis-Lung (MISCAN-Lung) framework to simulate the underlying lung cancer natural history, incorporating both preclinical and clinical phases under the Singapore setting (Figure 1).^25^ In the preclinical phase, carcinogenesis was assumed to occur after age 35 due to minimal incidence recorded prior to this age in Singapore, with the risk determined by smoking history, family history, age, and gender. Following carcinogenesis, cancers progressed sequentially through four stages (I–IV) with stage-specific, Weibull-distributed sojourn times.^25^ Individuals would enter the clinical phase upon diagnosis at the end of Stage I–III, or during Stage IV, with stage-specific probabilities. All individuals who developed lung cancer were assumed to be diagnosed unless death from other causes occurred first. In the clinical phase, no further stage progression was modelled, and individuals either died from lung cancer with stage-specific time-dependent hazard or survived to die later from other causes with stage-specific and time-dependent hazard. See the Supplementary Information for details on model configuration, parameter estimation, and validation.

**Figure 1.**
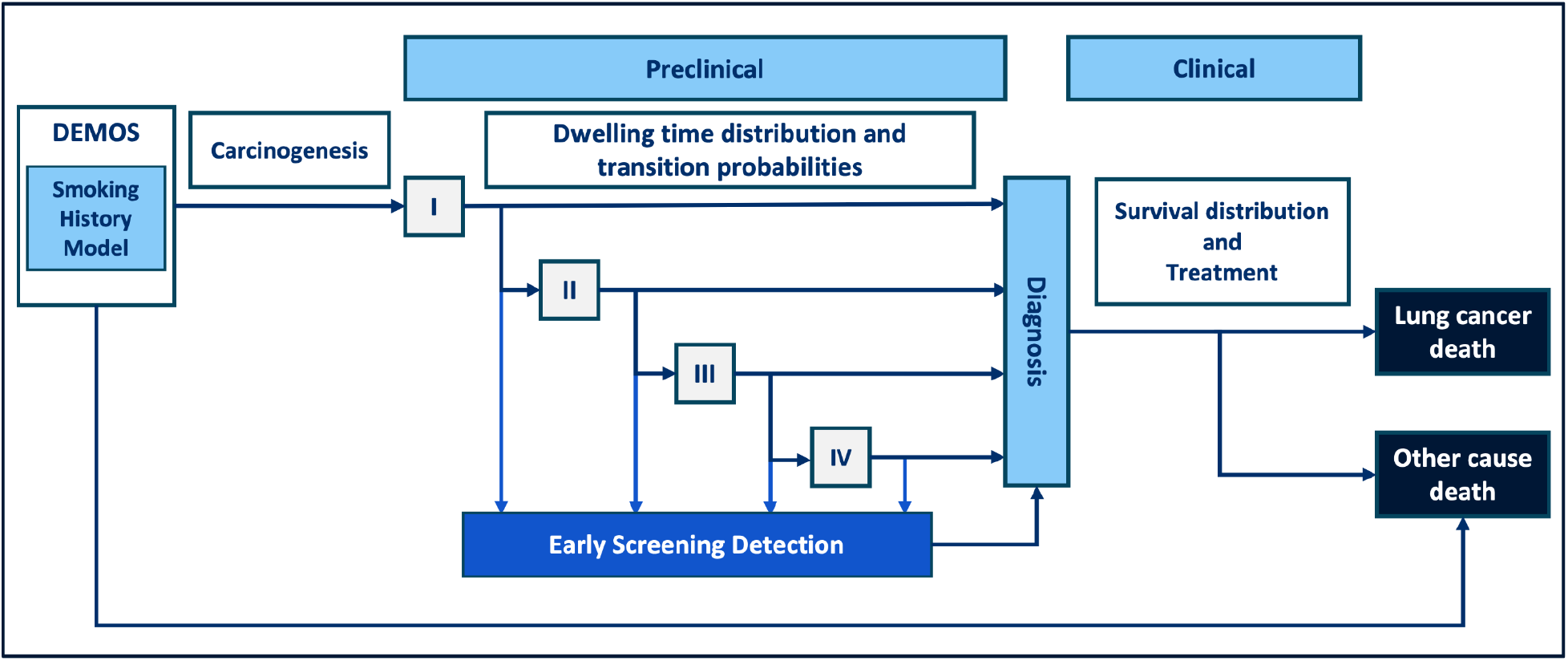
Model schematic.

### 2.5 Screening model

LDCT was selected for the lung cancer screening given its validated accuracy in detecting early-stage lung cancer and broad availability in Singapore.^26^ Individuals in the preclinical phase with a positive screening result were diagnosed at the screen-detected stage, while those with negative results remained in the preclinical phase until the next screen or symptomatic detection. Stage-specific screening sensitivity and overall specificity were applied (Table S4).

No screening was assumed during calibration due to the lack of historical screening uptake data. In projections, screening was implemented from 2025 to 2050 for male and female ever-smokers (≥20 pack-years for current smokers and ≤15 years since quitting for former smokers), reflecting their elevated risk and current guidelines.^13,27^ Eligibility began at ages 50, 55, or 60 years and continued until ages 75, 80, or 85 years, diagnosis, or death. Screening frequency ranged from annual to quinquennial, with uptake rates (proportion of eligible individuals screened in each round) of 40%, 70%, and 100%.

### 2.6 Economic model

Health benefits were assessed by reductions in late-stage incidence and deaths, and QALYs gained, relative to no screening within each tobacco control scenario. Costs were estimated from a healthcare-payer perspective, including screening and treatment, expressed in 2024 Singapore dollars (SGD) and discounted at 3% annually, with incremental costs and incremental cost-effectiveness ratios (ICERs) calculated relative to the no-additional-screening scenario. Together with overdiagnosis and false-positive rates, these metrics were used to identify the most life-saving with ≤10% overdiagnosis rate (MLSOD10), the most cost-effective, the lowest-overdiagnosis, and the overall optimal (TRS) screening strategies under each tobacco control scenario. Details on QALY, cost, and score calculations are provided in the Supplementary Information.

For each combination of tobacco control scenario and screening strategy, we ran 100 simulations of individual life trajectories to derive point estimates and 95% uncertainty intervals (UI) for the aforementioned metrics. Sensitivity analyses examined assumptions on survival extrapolation, test sensitivity, eligibility criteria (pack-years and time since quitting), and inclusion of individuals with a family history. All the analyses were performed in R (v4.5.3).^28^

## 3. Results

### 3.1 Projected smoking prevalence and disease burden without additional screening interventions

Under the status quo smoking scenario, smoking prevalence was projected to decrease from 8.2% (95% UI: 6.4%– 10.1%) in 2025 to 3.4% (95% UI: 1.6%–5.7%) by 2050, largely driven by cohort replacement. However, reductions in lung cancer burden decline were delayed, with annual incidence and mortality peaking at 2,824 (95% UI: 2,265– 3,715) and 1,732 (95% UI: 1,340–2,289) in 2040 and 2036, respectively, before declining to 2,579 (95% UI: 1,988– 3,572) and 1,267 (95% UI: 953–1,691) by 2050 (Figure 2). With tobacco control, the smoking prevalence was projected to decrease to 2.8% (95% UI: 1.3%–4.7%), 1.8% (95% UI: 0.75%–3.1%), 0.6% (95% UI: 0.2%–1.2%), and 0 for mild, moderate, stringent, and complete ban scenarios by 2050, respectively. Correspondingly, lung cancer incidence would reduce by 3.2% (95% UI: 2.4%–4.2%), 8.9% (95% UI: 6.8%–11.2%), 17.6% (95% UI: 14.0%– 21.7%), and 25.6% (95% UI: 20.6%–32.4%), while mortality would decrease by 3.2% (95% UI: 2.3%–4.1%), 9.1% (95% UI: 7.2%–11.5%), 18.6% (95% UI: 15.1%–22.4%), and 28.8% (95% UI: 23.2%–34.6%) by 2050, with the majority of these reductions attributable to males (Figure 2).

**Figure 2.**
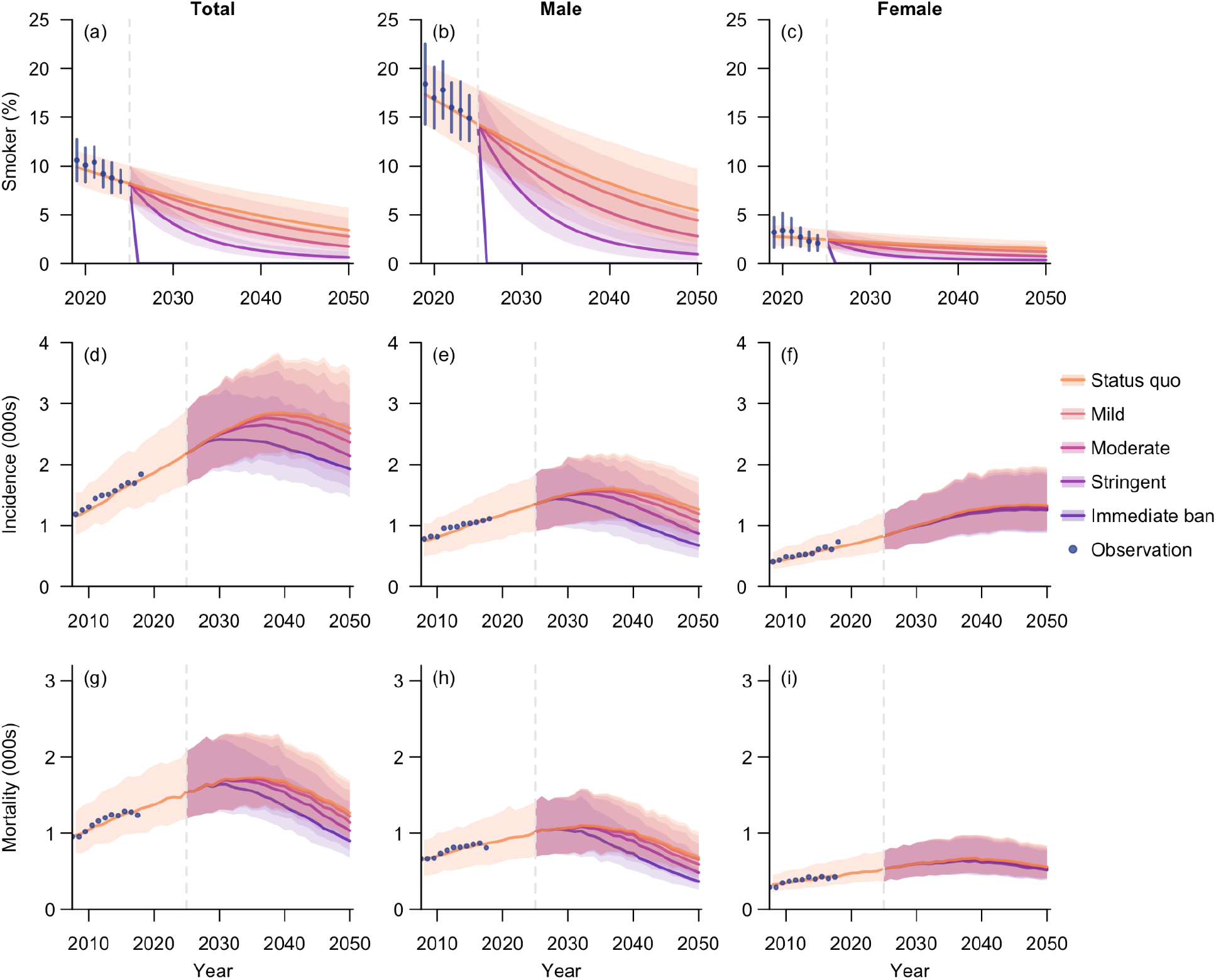
Model fits and baseline projections under five tobacco control scenarios. Row 1 shows projected smoking prevalence from 2019 to 2050, while Rows 2–3 show lung cancer incidence and mortality, respectively, from 2008 to 2050. Results are presented for the total population (Column 1) and stratified by gender (Columns 2– 3). Tobacco control campaigns were assumed to initiate in 2025, without screening implemented in any scenario. Projected means are represented by lines, with shades indicating the corresponding 95% uncertainty intervals. Observed values prior to 2025 were represented by blue dots, with vertical lines denoting 95% confidence intervals where available. Note the different time scales used in Row 1 and Rows 2–3.

### 3.2 Status quo with lung cancer screening

All evaluated strategies remained cost-effective under a willingness-to-pay threshold equal to the 2024 GDP per capita (∼SGD 120,000). Among strategies with ≤10% overdiagnosis, MLSOD10 was annual screening of all eligible ever-smokers aged 50–80 years, yielding 51,312 (95% UI: 36,821–72,830) QALYs at an additional cost of SGD 2.3 (95% UI: 1.9–2.7) billion versus no screening. TRS recommended biennial screening of ever-smokers aged 60–80 years, with lower QALY gains (32,861, 95% UI: 23,347–47,388) and costs (SGD 0.9, 95% UI: 0.7–1.2 billion), but substantially greater cost-effectiveness (ICER: 37,080 SGD/QALY, 95% UI: 27,826–48,774; Figure 3, Table S6–S7).

**Figure 3.**
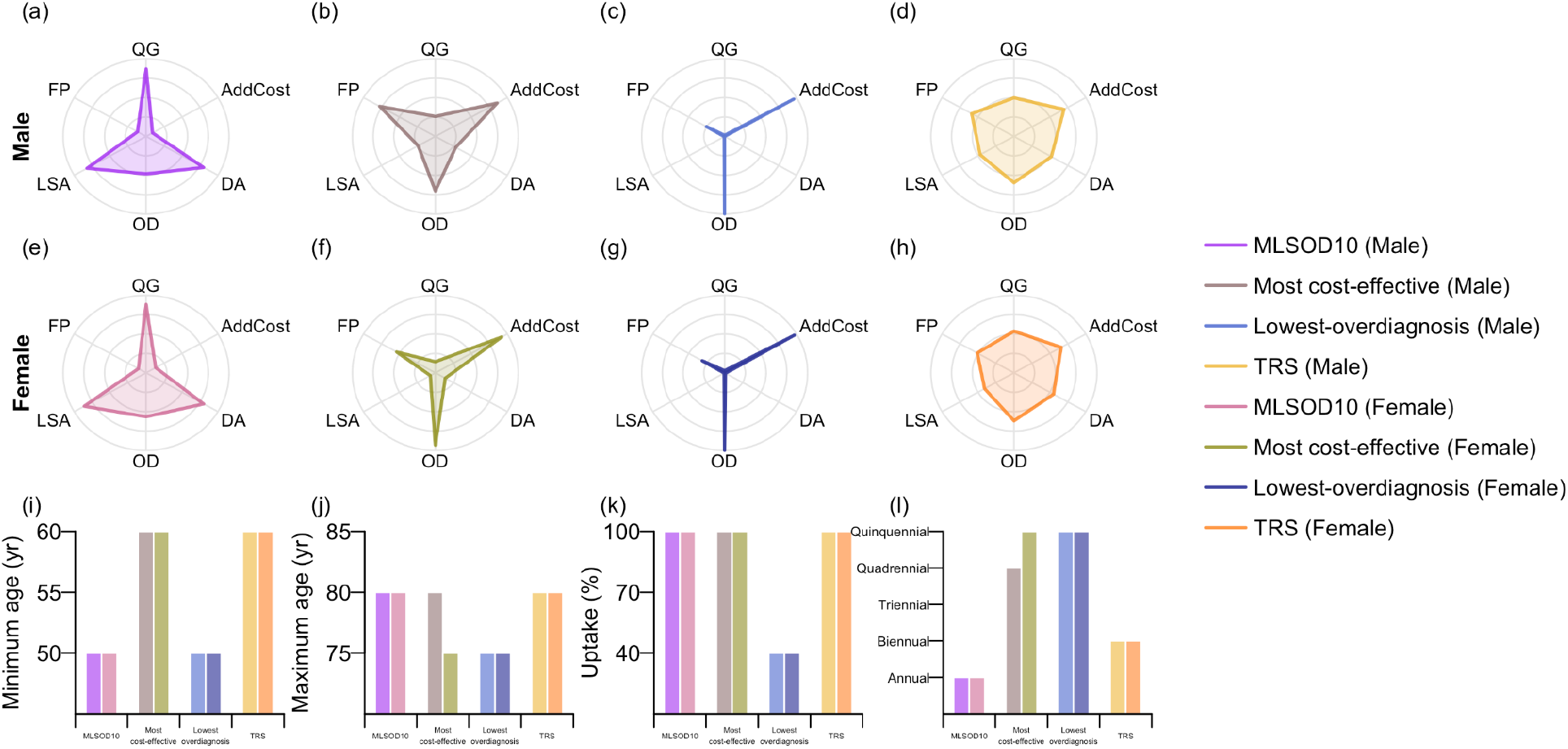
Recommended screening strategies under four decision criteria for the status quo scenario. The four selected strategies comprise the most life-saving with ≤10% overdiagnosis rate (MLSOD10), the most cost-effective, the lowest-overdiagnosis, and the overall optimal strategy (TRS). Subfigures (a)–(h) (Row 1 for male ever-smokers and Row 2 for female ever-smokers) present the scores for six metrics evaluated over year 2025– 2050: QALY gains (QG), late-stage cancer averted (LSA), deaths averted (DA), and additional costs (AddCost) relative to the scenario without additional screening, as well as overdiagnosis rate (OD) and false positive rate (FP). All the metrics represent medians across simulations and are rescaled to a 0–1 range. Subfigures (i)–(l) (Row 3) show the characteristics of the selected strategies, including minimum age, maximum age, screening uptake rate, and screening frequency (rescaled to an annual rate). Strategies selected under different criteria for male and female ever-smokers are distinguished by colour, as indicated in the legend on the right.

Compared to MLSOD10, reducing the maximum eligible age to 75 years decreased QALY gains by 21.3% (95% UI: 12.9%–23.6%) and incremental costs by 12.9% (95% UI: 10.5%–16.1%), increasing the ICER by 10.3% (95% UI: 8.5%–12.2%). Raising the minimum eligible age to 55 years reduced QALY gains and additional costs by 3.8% (95% UI: 2.5%–5.2%) and 13.2% (95% UI: 9.1%–16.7%), respectively, lowering the ICER by 9.7% (95% UI: 6.9%–12.3%), indicating higher cost-effectiveness when targeting older populations. Lower uptake and less frequent screening further reduced QALYs, costs, and ICERs, with ICERs decreasing by 23.1% (95% UI: 20.6%–25.3%) at 40% uptake and 30.2% (95% UI: 26.3%–34.3%) under quinquennial screening. Overdiagnosis rate increased by 45.5% (95% UI: 41.5%–49.8%) with a 5-year increase in the maximum eligible age but changed little with the minimum age. It declined by 18.0% (95% UI: 14.9%–20.7%) at 40% uptake and 29.7% (95% UI: 26.1%–33.7%) with quinquennial screening. False-positive rates changed minimally (<2%; Figure 4). These patterns were consistent across gender and alternative reference strategies, though with varying magnitudes (Figure S6).

**Figure 4.**
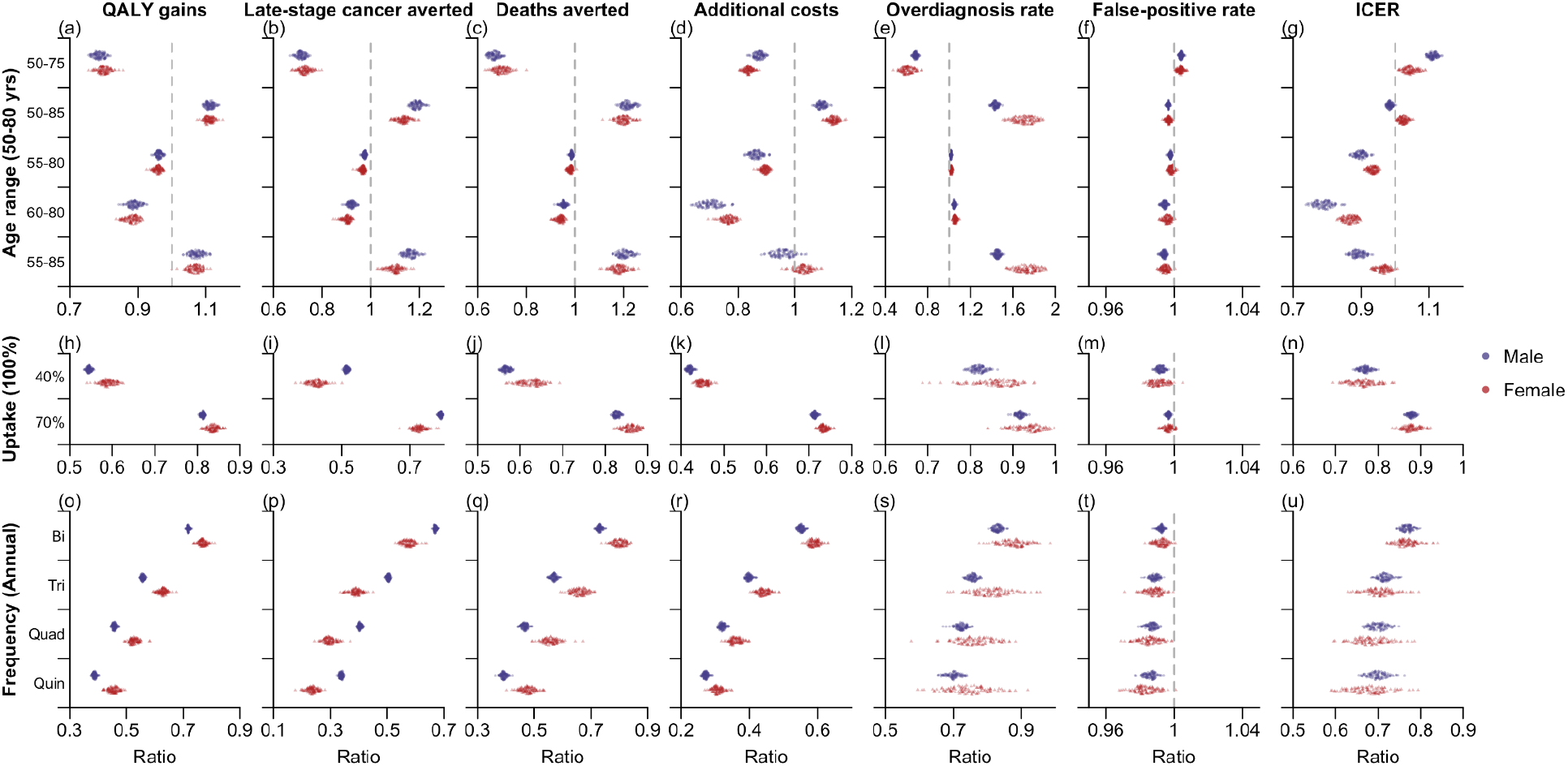
Comparison of MLSOD10 with alternative screening strategies. Ratios of outcomes for each alternative strategy relative to MLSOD10 (denominator; annual screening of eligible ever-smokers aged 50–80 years) were computed per simulation and shown as dots, with blue and red indicating males and females, respectively. Each alternative strategy differs from MLSOD10 in one attribute (age range, uptake, or screening frequency), with reference values given in parentheses on the left and alternative values listed on the y-axis. All strategies were evaluated over 2025–2050 across seven metrics (Columns 1–7): QALY gains, late-stage cancers averted, deaths averted, additional costs, overdiagnosis rate, false-positive rate, and ICER, each relative to the scenario without additional screening.

### 3.3 Smoking ban with lung cancer screening

The effects of screening strategy attributes on marginal health and economic outcomes were largely consistent across tobacco control scenarios, but tobacco control had a substantially larger impact than screening alone (Figure 5). Compared with their status quo counterparts, implementing MLSOD10 alongside an immediate smoking ban yielded an additional 146,924 QALYs (95% UI: 95,732–203,464), 40,570 fewer late-stage cases (95% UI: 23,851– 59,643), and 24,556 fewer deaths (95% UI: 16,183–34,978), while reducing total costs by 2.9 (95% UI: 2.0–4.0) billion, with 1,226 (95% UI: 848–1,795) fewer overdiagnosed cases and 302,564 (95% UI: 225,250–365,443) false-positive cases. Similarly, under the immediate ban scenario, TRS increased QALYs by 154,984 (95% UI: 101,097– 214,557) and reduced total costs by 2.5 (95% UI: 1.7–3.5) billion relative to TRS under the status quo. Despite a moderate increase in ICER (by ∼14,000 SGD/QALY), both MLSOD10 and TRS remained cost-effective under the immediate ban scenario.

**Figure 5.**
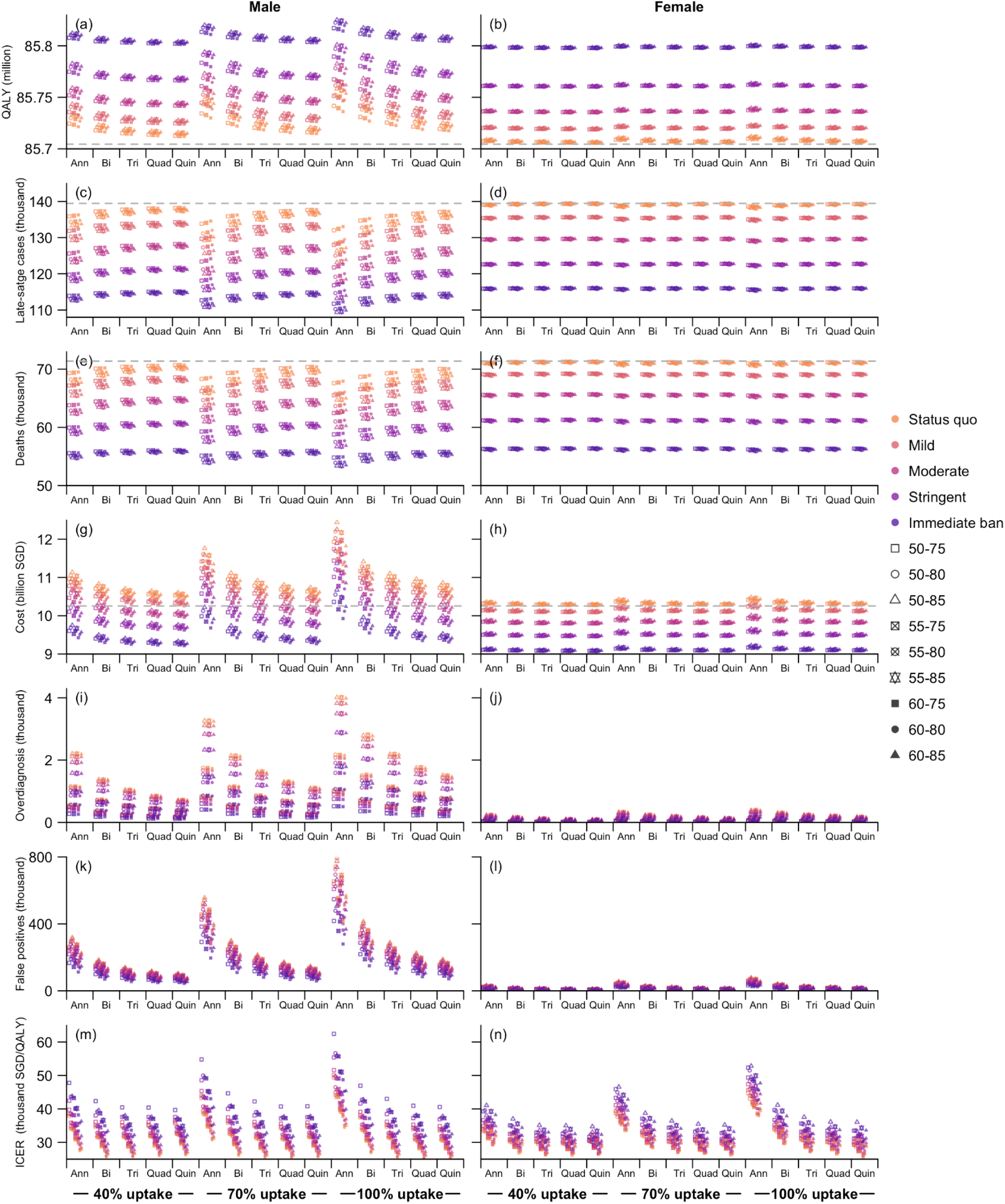
Comparison of screening strategies across five tobacco control scenarios. All strategies were evaluated over 2025–2050 using seven metrics (Rows 1–7): total QALYs, late-stage cancers, deaths, costs, overdiagnosed cases, false-positives cases, and ICER. Points show median estimates across simulations for each strategy. Strategies are defined by target age range, screening frequencies, and uptake, with frequency-uptake combinations shown on the x-axis and age range indicated by different point styles (see legend). The grey vertical lines in subfigures (a)–(h) (Rows 1–4) denote the corresponding values under the status quo scenario without screening. Note that ICERs in subfigure (m)–(n) were for screening only, calculated relative to no additional screening within each tobacco control setting.

Screening could further be extended to other at-risk populations. Expanding MLSOD10 to ever-smokers while relaxing eligibility to ≥1 pack-year for current smokers and ≤20 years since quitting for former smokers remained cost-effective for both genders, with ICERs up to 102,159 (95% UI: 66,229–135,507) SGD/QALY (Figure S13). Screening individuals with a first-degree family history of lung cancer alone was cost-effective only for females aged ≥55 years at low frequencies (triennial or less; Figure S17–S18), but became cost-effective up to biennial when combined with MLSOD10 or TRS for ever-smokers (Figure S15–S16). Applying MLSOD10 to ever-smokers and extending to the remaining population with a family history (also identified as MLSOD10 in this group) yielded QALY gains of 746 (95% UI: 181–2,457) at an incremental cost of SGD 112 (95% UI: 43–448) million compared to screening eligible ever-smokers alone (Figure S17, S19).

## 4. Discussion

In this study, we constructed a national microsimulation model to evaluate the long-term health and economic impacts of lung cancer screening strategies under alternative tobacco control scenarios, including a national complete ban, in Singapore from 2025 to 2050. By explicitly simulating individual life histories, smoking trajectories, and disease progression, the model captured heterogeneity in risk profiles and allowed joint assessment of downstream clinical interventions and upstream prevention within a unified framework.

Compared to MLSOD10 under status quo smoking projections, implementing an immediate smoking ban yielded 282% (95% UI: 220%–346%) more QALYs gained and 432% (95% UI: 325%–564%) more deaths averted, with corresponding figures under TRS of 460% (95% UI: 347%–578%) and 612% (95% UI: 444%–825%) respectively. In absolute terms, the ban produced similar reductions for both MLSOD10 and TRS where deaths fell by 39% (95% UI: 29%–50%) and total costs by 23% (95% UI: 17%–30%). The magnitude of benefit observed under combined ban and screening scenarios is consistent with, and extends beyond, trial-level evidence on the interaction between smoking abstinence and LDCT. Within the National Lung Screening Trial, sustained smoking abstinence over seven years reduced lung cancer mortality by 20%, equivalent to approximately three annual rounds of LDCT screening, with combined effects yielding a 38% reduction in lung cancer mortality (HR 0.62, 95% CI: 0.51–0.76).^11^ In the Multicentric Italian Lung Detection Trial, cessation both prior to screening (HR 0.57, 95% CI: 0.38–0.85) and during follow-up (HR 0.65, 95% CI: 0.44–0.96) was associated with significant mortality reductions.^29^ Furthermore, the CISNET MichiganLung model analysis of the 1960 US birth cohort demonstrated that a one-time cessation intervention of just 15% effectiveness at the point of screening increased deaths averted by approximately 36% and life-years gained by approximately 207% compared to without.^30^ Critically, a cessation intervention of this modest effectiveness produced life-years gained comparable to maximising screening uptake from 30% to 100%.^31^

Alongside the implementation of a smoking ban, we found that both MLSOD10 and TRS strategies were largely cost-effective, despite the expected diminishing high-risk smoking population. We further found that expanding the screening eligible population to ever-smokers that had quit within 20 years, and those with family history for all age ranges still yielded cost-effective evaluations. Similar findings regarding the cost-effectiveness of screening individuals with older smoking history or first-degree family history (with assumed familial clustering of shared genetic susceptibility or shared environmental smoke exposure within household exposure), and at younger age ranges, have been observed.^32–35^ Whilst alternative approaches, including polygenic risk scoring, have shown promise for improving lung cancer risk stratification with genome-wide association studies identifying susceptibility loci such as 15q25, 5p15, and 6p21, their clinical utility is currently constrained by limited genome-wide genotyping data availability and modest predictive gains over established factors.^36,37^ Regardless, risk stratification is likely to improve with increasing genetic data availability, and among individuals at higher risk, our analysis suggests that LDCT screening should continue even in the context of complete and permanent smoking cessation.

Singapore represents one of the few settings globally in which a complete tobacco ban constitutes a plausible near-term policy scenario. Several challenges however require acknowledgement. Bhutan, which enacted the world’s first national tobacco sales prohibition in 2004, found that overall tobacco use rose from 18.5% in 2004 to 27.3% in 2019, despite strict enforcement including long prison sentences, due to illegal smuggling.^38,39^ The ban was partially reversed under the Tobacco Control Act 2021, effectively acknowledging that supply side prohibition had failed to eliminate demand.^40^ Similarly, New Zealand’s smokefree generation legislation, enacted in December, was eventually repealed in February 2024.^41,42^ The IARC modelling study itself noted that compliance with analogous age-based tobacco restrictions has varied between 62% and 94% in practice, and recommended that generational bans be implemented alongside proven demand side measures including taxation, cessation support, and smoke-free environments.^43^ Singapore differs from both settings in its already low smoking prevalence of 8.4%, absolute bans for smoking in public spaces, stringent border control, high institutional trust, and a regulatory environment that has already banned vaping with very low non-compliance. A substantial body of evidence exists on the benefits of smoking cessation on population health, where lung cancer burden reduction, with further screening support for high-risk groups, could greatly alleviate healthcare system strain and costs.^43–45^

In a wider context, MLSOD10 could be recommended for high income settings and TRS for lower income settings due to the reduced screening demand of the latter, and its estimated higher cost-effectiveness. Whilst MLSOD10 focuses on maximum life gains, considering overdiagnosis in its trade off, TRS balances health gains with costs. Alternative ICER-based decision rules likely favour strategies with modest health benefits, while completely health-outcome-based approaches, which included more lower-risk groups, may incur substantial overdiagnosis and economic burden, highlighting the limitations of relying on a single outcome.^46^ The two approaches presented here accommodate the nuances that both clinicians and policymakers consider in recommending lung cancer screening to large populations, and further demonstrate the contribution of smoking bans with LDCT screening in place.

Our study has limitations. Firstly, the model only included smoking dose and family history as risk factors, omitting other exposures such as passive or occupational smoke exposure.^47–49^ This omission restricted the evaluation of targeted strategies for individuals with such exposures, and might have underestimated the spillover effects of tobacco control. Another limitation of this study pertains to the available training data. Smoking prevalence calibration relied on self-reported data, potentially underestimating prevalence and potentially overestimating non-smoker carcinogenesis risk. Additionally, although immigration dynamics were incorporated, smoking behaviours among immigrants were assumed to be similar to those of local residents. In the absence of relevant data, the model furthermore did not distinguish between molecular subtypes, despite well-documented heterogeneity in histology, prognosis, and survival across lung cancer subtypes.^25^ We also assumed homogeneous tumour progression rates, treatment responses and associated costs, which may further limit precision. New screening technologies and risk stratification criteria determining individuals at high risk could also affect our projected screening evaluations.

Despite the limitations, our microsimulation framework enables individual-level life histories to be generated, capturing the pathway from smoking through carcinogenesis, disease progression, detection, and outcomes, with transition risks dependent on prior exposures and timing. This approach extends beyond conventional cohort-based Markov models, facilitating the health and economic evaluation of targeted screening and tobacco control interventions, both independently and in combination. Multiple simulations were also performed to capture parameter uncertainty and stochasticity in individual-level events, enhancing projection credibility. In addition, the framework developed is readily adaptable to other settings to evaluate the joint impact of population ageing, evolving smoking behaviours, screening, and tobacco control.

Singapore is among the few settings in which tobacco elimination may be viewed as a credible long term public health ambition, supported by low smoking prevalence and a strong history of regulatory action. While a full immediate ban would be challenging to implement and would require careful attention to enforcement, compliance, and unintended effects, the projected magnitude of benefit indicates that tobacco elimination should be part of the policy conversation. The complete ban scenario therefore provides a useful benchmark for understanding the potential gains from ambitious endgame policies and for motivating discussion of feasible pathways towards elimination in Singapore and comparable settings.

## Data Availability

Source data utilised for this study are available upon request.

## Contributors

YH, SJ, and BLD conceived and designed the study. YH implemented the statistical analysis. YH and SJ created the figures and tables. SJ wrote the original draft of the manuscript. YH, SJ, XZ, KIF, YW, KBT, RS, JTL, and BLD reviewed and edited the manuscript.

## Data sharing statement

Source data utilised for this study are available upon request. Analytical scripts are available at https://github.com/Mister-He/DEMOS-MISCAN-Lung-Public.

## Acknowledgments

This work was supported by the National Medical Research Council (NMRC) Population Health Research Grant - New Investigator Grant (MOH-001430-00 PHRG-NIG) and Population Health Metrics and Analytics project, funded by the Ministry of Health, Singapore.

