## Supplementary Information for "Time for Tobacco Elimination: Modelling smoking cessation strategies and lung cancer screening in Singapore"

### Data sources

#### Multi-Ethnic Cohort Study

The de-identified longitudinal smoking history data utilised to calibrate the smoking history model were sourced from the Singapore Multi-Ethnic Cohort Phase 1 (MEC1),[1] a closed cohort comprising 14,600 adults aged 20 years and above. MEC1 was established by combining participants from the Singapore Prospective Study (SP2) and the Singapore Cardiovascular Cohort Study (SCCS2), conducted between 2004 and 2007. An additional 6,300 participants were recruited during 2007–2010, comprising SCCS-MEC. Data from the follow-up study conducted in 2011–2016 were also used for calibration.

The cohort is representative of Singapore's three major ethnic groups: Chinese, Malay and Indian. Participants were originally drawn from four cross-sectional surveys: the Thyroid and Heart Study (1982–1984), the National Health Survey (1992, 1998), and the National University of Singapore Heart Study (1993–1995). Ethical approval was obtained from the SingHealth Centralised Institutional Review Board for SP2 and the National University of Singapore Institutional Review Board for the remaining MEC1 studies.

#### Singapore Cancer Registry

Population-level lung cancer data, including five-year incidence and mortality counts stratified by five-year age group, gender, ethnicity, and stage (where applicable), as well as stage-specific survival probabilities, were aggregated from de-identified individual-level records from the Singapore Cancer Registry, provided by the National Registry of Diseases Office (NRDO).[2] The registry captures nearly all diagnosed lung cancer cases (n = 20,733) among Singapore residents and their associated deaths from both public and private hospitals between 1968 and 2023.

Beginning in 2003, individual patient records also included the lung cancer stage at diagnosis alongside the date of diagnosis. Stages for cases diagnosed between 2003 and 2018 were defined according to the American Joint Committee on Cancer (AJCC) 6th and 7th editions, based on anatomical staging, while those diagnosed from 2018 onwards followed the AJCC 8th edition, based on prognostic staging. As the two systems are not directly comparable, cases staged under the 8th edition were analyzed separately. The data indicated a moderate shift towards earlier detection, with the proportion of Stage I–II diagnoses increasing from 13hao8% in 2003–2007 to 23.2% in 2013–2017.

#### Singapore Translational Cancer Consortium

Parameters for the natural history model and treatment cost model were estimated using data from Singapore Translational Cancer Consortium (STCC),[3] which include annual incidence and death counts by age and gender, as well as direct medical expenditures associated with lung cancer treatment and care from 2009 to 2019. Costs were aggregated by smoking status (never- and ever-smoker), diagnosed stage (I–IV), and time since diagnosis (1–10 years), with the corresponding sample size also reported for each stratum.

### Methods

#### Smoking history model

#### Rationale for model structure and parameterization

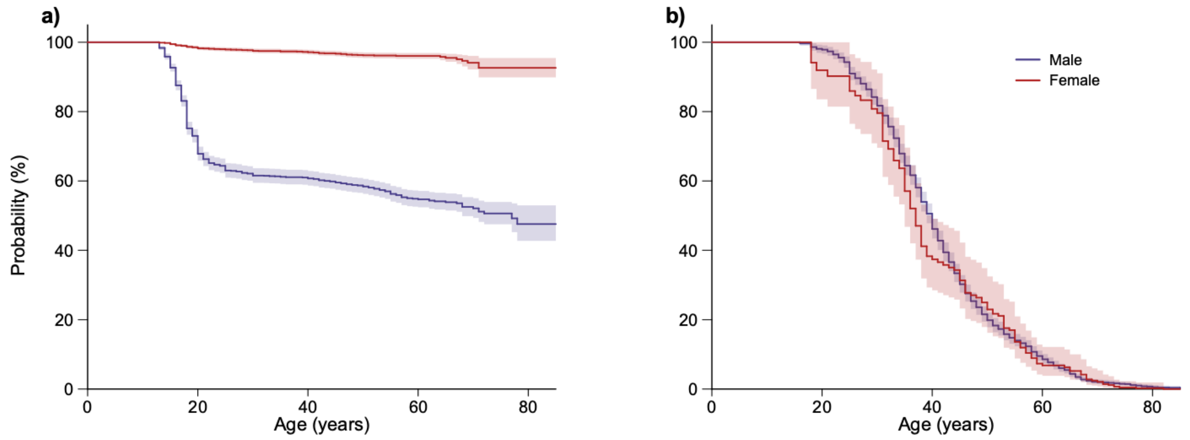

**Number at risk**

| | 0 | 20 | 40 | 60 | 80 |
|---|---|---|---|---|---|
| Male | 2 074 | 1 514 | 1 181 | 335 | 15 |
| Female | 2 339 | 2 305 | 2 185 | 572 | 13 |

**Number at risk**

| | 0 | 20 | 40 | 60 | 80 |
|---|---|---|---|---|---|
| Male | 301 | 838 | 651 | 163 | 8 |
| Female | 34 | 54 | 42 | 14 | 1 |

**Figure S1. Gender- and age- specific probabilities of a) remaining never-smokers in the overall population and (b) remaining current smokers among ever-smokers, estimated from the MEC1 data.** Mean estimates are shown as lines, with the corresponding 95% CIs indicated by shaded areas. Number at risk tables are listed below subfigures.

We derived the gender- and age- specific survival probabilities for remaining never-smokers in the overall population and for not quitting among ever-smokers, based on self-reported values in the MEC1 data (Figure S1). The estimates suggest that majority of the male ever-smokers initiated between ages 15 and 30, with the highest probabilities between 18 and 25 years. In contrast, smoking initiation among females was much lower, despite a slight increase after age 50. Overall, the results suggest a unimodal pattern of smoking initiation across the life span, starting from approximately zero before age 15, peaking around age 20, and waning gradually towards zero by age 40. Regarding smoking cessation, both males and females exhibited an approximately constant hazard of quitting across the adult age range (20–79 years), indicating that cessation hazard is largely age-invariant.

#### Model specification

The transition probabilities between smoking states were modelled as functions of age, gender, ethnicity, and birth cohort. The birth cohort was defined based on the individual's year of birth, grouped into five-year intervals (e.g., 1970–1974, 1975–1979), and coded as an integer-valued variable, with the reference group (0) corresponding to those born in 1970–1974. For an individual of gender $g_i$ , ethnicity $r_i$ and birth cohort $b_i$, the probability of initiating smoking at $a_i$ years old, given that this individual was previously a never smoker, was

$$p_{i,1}(a_i) = 1 - \exp(-\exp(\beta_{0,1} + \gamma_1(g_i, r_i) + \beta_{1,1} \cdot b_i - \beta_{2,1}(a_i - 20)^2)),$$

where $\beta_{0,1}$ is the intercept, $\gamma_1$ captures the combined impact of gender and ethnicity, $\beta_{1,1}$ represents the influence of birth cohort, and $\beta_{2,1} > 0$ indicates the bell-shape impact of age with the initiation hazard peak at 20 years old. Similarly, if the individual was a current smoker, the probability of quitting at age $a_i$ years was

$$p_{i,2}(a_i) = 1 - \exp(-\exp(\beta_{0,2} + \gamma_2(g_i, r_i) + \beta_{1,2} \cdot b_i + \beta_{2,2}(a_i - 40))),$$

where the coefficient $\beta_{2,2} > 0$ ensures that the cessation probability increases monotonically with age.

We first estimated the parameters ($\beta_{0:2,1:2}$ and $\gamma_{1:2}$) within a Bayesian framework using self-reported smoking history data from the MEC1,[1] employing Hamiltonian Monte Carlo (HMC) implemented via the No-U-Turn Sampler (NUTS) in Stan (accessed through R). Four parallel Markov chains were run, each with 2,000 warm-up iterations followed by 2,000 sampling iterations, yielding a total of 8,000 posterior samples. Convergence was assessed through visual inspection of trace plots, effective sample sizes (>1,000), and the Gelman–Rubin diagnostic (R-hat <1.01).

To minimise impact of cohort sampling bias on inference, we adjusted the posterior predictive distribution of the initiation and cessation probabilities by applying gender- and cohort-specific scaling factors $s_k(g,b) = \lambda_{g,k} + \psi_{g,k} \cdot b$ to obtain the adjusted transition probabilities from never smoker to current smoker ($k = 1$)and from current smoker to former smoker ($k = 2$).

Values of $\lambda_{g,1:2}$ and $\psi_{g,1:2}$ were informed by the gender-specific smoking prevalence estimates reported annually from 2019 to 2024 in the National Population Health Surveys.[4] Let $q_{g,y}$ denote the point estimate of smoking prevalence for gender $g$ and year $y$, and $[l_{g,y}, u_{g,y}]$ represent the corresponding 95% confidence interval (CI). We minimised the following objective function on the logit scale:

$$L(\lambda,\theta) = \sum_g \sum_{y=2019}^{2024} (\text{logit}(\hat{p}_{g,y}) - \text{logit}(p_{g,y}))^2 / (((\text{logit}(u_{g,y}) - \text{logit}(l_{g,y}))/3.84)^2,$$

where $\hat{p}_{g,y}$ is the projected smoking prevalence for gender $g$ and year $y$ generated by DEMOS using each posterior draw of the model parameters and scaled by the gender- and cohort- specific scaling factors determined through $\lambda_g$ and $\theta_g$. All model parameters and data sources are provided in Table S1.

**Table S1. Parameters of the smoking history model and natural history model with corresponding values or prior distributions.**

| Parameter | Definition | Source and reported/estimated value (with 95% credible interval or CI, where applicable) |
|---|---|---|
| $\beta_{0,1}$ | Intercept of smoking initiation hazard | Estimated; -3.32 (-3.51– -3.13) |
| $\beta_{1,1}$ | Birth cohort effect on smoking initiation hazard | Estimated; 0.045 (0.027–0.068) |
| $\beta_{2,1}$ | Age effect on smoking initiation hazard | Estimated; 0.014 (0.013–0.015) |
| $\gamma_1$ | Combined effect of gender and ethnicity on smoking initiation hazard | Estimated;<br>Male Chinese: 0.023 (95% UI: 0.019–0.027)<br>Male Malay: 0.79 (95% UI: 0.78–0.79)<br>Male Indian: 0.30 (95% UI: 0.30–0.31)<br>Male Other: 0.48 (95% UI: 0.48–0.49)<br>Female Chinese: -2.59 (95% UI: -2.60– -2.57)<br>Female Malay: -2.34 (95% UI: -2.36– -2.33)<br>Female Indian: -3.24 (95% UI: -3.26– -3.22)<br>Female Other: -2.48 (95% UI: -2.49– -2.46) |
| $\beta_{0,2}$ | Intercept of smoking cessation hazard | Estimated; -3.28 (-3.48– -3.08) |
| $\beta_{1,2}$ | Birth cohort effect on smoking cessation hazard | Estimated; 0.14 (0.11–0.17) |
| $\beta_{2,2}$ | Age effect on smoking cessation hazard | Estimated; 0.003 (0.002–0.007) |
| $\gamma_2$ | Combined effect of gender and ethnicity on smoking cessation hazard | Estimated;<br>Male Chinese: 1.57 (95% UI: 1.56–1.58)<br>Male Malay: 1.70 (95% UI: 1.68–1.71)<br>Male Indian: 1.59 (95% UI: 1.58–1.60)<br>Male Other: 1.72 (95% UI: 1.71–1.73)<br>Female Chinese: 1.65 (95% UI: 1.64–1.66)<br>Female Malay: 1.47 (95% UI: 1.46–1.49)<br>Female Indian: 1.08 (95% UI: 1.07–1.09)<br>Female Other: 1.10 (95% UI: 1.09–1.11) |
| $\lambda_{male,1}$ | Intercept of the scaling factor for male smoking initiation | Estimated; -0.18 (-0.31–0.07) |
| $\psi_{male,1}$ | Birth cohort effect on the scaling factor for male smoking initiation | Estimated; 1.48 (1.08–1.85) |
| $\lambda_{female,1}$ | Intercept of the scaling factor for female smoking initiation | Estimated; 0.33 (0.09–0.56) |
| $\psi_{female,1}$ | Birth cohort effect on the scaling factor for female smoking initiation | Estimated; -0.31 (-0.85– -0.14) |
| $\lambda_{male,2}$ | Intercept of the scaling factor for male smoking cessation | Estimated; -1.79 (-1.88– -1.65) |

| | | |
|---|---|---|
| $\psi_{male,2}$ | Birth cohort effect on the scaling factor for male smoking cessation | Estimated; -1.89 (-2.60– -1.76) |
| $\lambda_{female,2}$ | Intercept of the scaling factor for female smoking cessation | Estimated; -1.32 (-1.57– -1.16) |
| $\psi_{female,2}$ | Birth cohort effect on the scaling factor for female smoking cessation | Estimated; -1.09 (-1.33– -0.54) |
| $\alpha_{0,male}$ | Intercept for baseline never-smoker male carcinogenesis hazard | Estimated; -14.439 (-14.442– -14.437) |
| $\gamma_{male}$ | Age effect on baseline never-smoker male carcinogenesis hazard | Estimated; 0.092 (0.090–0.095) |
| $\alpha_{0,female}$ | Intercept for baseline never-smoker female carcinogenesis hazard | Estimated; -12.72 (-12.72– -12.71) |
| $\gamma_{female}$ | Age effect on baseline never-smoker female carcinogenesis hazard | Estimated; 0.084 (0.082–0.087) |
| $HR_f$ | Hazard ratio for carcinogenesis in individuals with versus without a first-degree family history | Kishida et al.;[5] 1.45 |
| $OR(py_i \| s_i = 1, g_i = M)$ | Odds ratios for carcinogenesis among male current smokers by pack-year level (reference: never-smoker) | Pesch et al.;[6]<br>1–<20: 8.9<br>20–<30: 17.1<br>30–<40: 24.6<br>40–<50: 32.4<br>50–<60: 46.3<br>≥60: 47.7 |
| $OR(py_i \| s_i = 1, g_i = F)$ | Odds ratios for carcinogenesis among female current smokers by pack-year level (reference: never-smoker) | Pesch et al.;[6]<br>1–<20: 3.5<br>20–<30: 17.1<br>30–<40: 12.9<br>40–<50: 32.4<br>50–<60: 46.3<br>≥60: 47.7 |
| $OR(qy_i \| s_i = 2, g_i = M)$ | Odds ratios for carcinogenesis among male former smokers by pack-year level (reference: never-smoker) | Pesch et al.;[6]<br><2: 23.6<br>2–5: 18.3<br>6–10: 10.8<br>11–15: 7.8<br>16–25: 5.1<br>26–35: 2.9<br>>35: 2.2 |
| $OR(qy_i \| s_i = 2, g_i = F)$ | Odds ratios for carcinogenesis among female former smokers by pack-year level (reference: never-smoker) | Pesch et al.;[6]<br><2: 7.8<br>2–5: 6.7<br>6–10: 4.0<br>11–15: 3.3<br>16–25: 2.0<br>26–35: 1.0<br>>35: 1.3 |
| $\theta_{0,k_i}$ | Stage-specific intercept for survival hazards | Estimated;<br>Stage I: -0.006 (-0.026– 0.015)<br>Stage II: -0.12 (-0.16– -0.09)<br>Stage III: -0.20 (-0.22– -0.18)<br>Stage IV: -0.21 (-0.22– -0.19) |
| $\theta_{1,k_i}$ | Stage-specific temporal effect on survival hazards | Estimated;<br>Stage I: -2.87 (-2.97– -2.77)<br>Stage II: -1.79 (-1.92– -1.67)<br>Stage III: -1.04 (-1.10– -0.98)<br>Stage IV: -0.44 (-0.47– -0.40) |
| $\theta_2$ | Impact of smoke status on survival hazards | Estimated; 0.57 (0.54–0.60) |
| $\theta_3$ | Impact of diagnosis year on survival hazards | Estimated; -0.033 (-0.037– -0.029) |

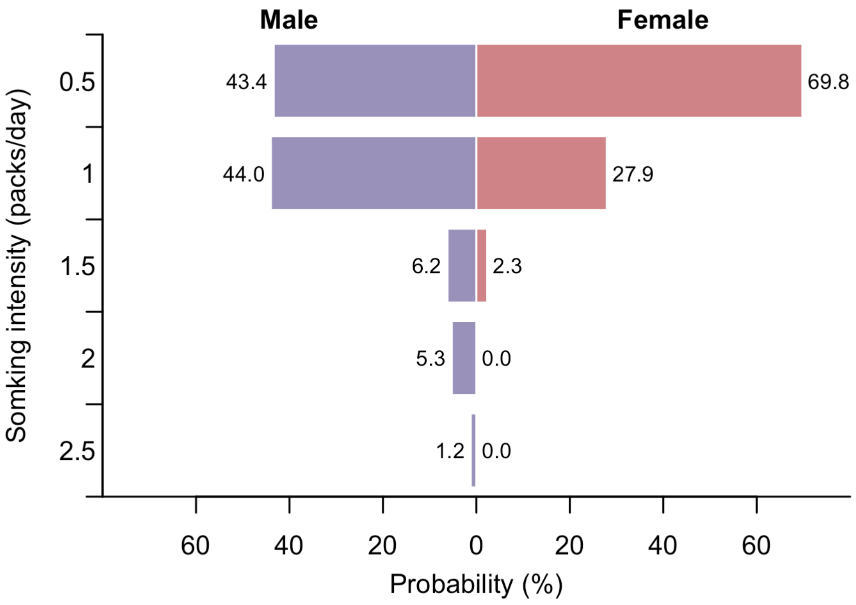

**Figure S2. Empirical distribution of smoking intensity, defined as the average number of cigarette packs smoked per day, among male (left) and female (right) ever-smokers.**

### Natural history model

#### Carcinogenesis

The preclinical phase began with the onset of carcinogenesis, which was assumed to occur only after age 35 due to the minimal incidence of lung cancer recorded in the Singapore Cancer Registry.[2] Individual smoking profiles generated by DEMOS were the only modelled risk factor. For a healthy never-smoker (smoking status $s_i = 0$) of gender $g_i$, the baseline hazard of carcinogenesis at age $a_i (> 35)$ was given by

$$h_{i,LC}(a_i \mid s_i = 0) = \exp\left(\alpha_{0,g_i} + \gamma_{g_i} a_i\right),$$

where $\alpha_{0,g_i}$ capture gender-specific effects, and $\gamma_{g_i} > 0$ represents the age effect.

The impact of first-degree family history of lung cancer, present in 3.5% of the population, was modelled using a fixed hazard ratio of 1.45 ($HR_f$).[5] Accordingly, the age-specific carcinogenesis hazard for individuals with a family history ($f_i = 1$) was specified as

$$h_{i,LC}(a_i \mid f_i = 1) = h_{i,LC}(a_i \mid f_i = 0) \cdot HR_f \,.$$

Assuming annual updates of carcinogenesis hazard, the annual probability of carcinogenesis for a never-smoker with family history status $f_i$ at age $a_i$ was calculated as

$$p_{i,LC}(a_i \mid f_i, s_i = 0) = 1 - \exp\left(-h_{i,LC}(a_i \mid f_i, s_i = 0)\right).$$

Smoking-related effects on carcinogenesis were accounted for by applying smoking- and gender- specific odds ratios. For current smokers ($s_i = 1$) of gender $g_i$, the odds ratio, $OR(py_i | s_i = 1, g_i)$, was a function of cumulative exposure, quantified by pack-years (i.e., smoking intensity multiplied by duration) grouped into six categories (1–<20, 20–<30, 30–<40, 40–<50,50–<60, ≥60; denoted as $py_i$), such that the corresponding carcinogenesis probability at age $a_i$ was

$$p_{i,LC}(a_i \mid f_i, s_i = 1) = \frac{p_{i,LC}(a_i \mid f_i, s_i = 0) \cdot OR(py_i \mid s_i = 1, g_i)}{1 + (OR(py_i | s_i = 1, g_i) - 1) \cdot p_{i,LC}(a_i \mid f_i, s_i = 0)}.$$

Similarly, for former smokers ($s_i = 2$) of gender $g_i$, the odds ratio, $OR(qy_i|s_i = 2, g_i)$, was determined by years since quitting classified into seven categories (<2, 2–5, 6–10, 11–15, 16–25, 26–35, and >35; denoted as $qy_i$), in the form of

$$p_{i,LC}(a_i \mid f_i, s_i = 2) = \frac{p_{i,LC}(a_i \mid f_i, s_i = 0) \cdot OR(qy_i \mid s_i = 2, g_i)}{1 + (OR(qy_i \mid s_i = 2, g_i) - 1) \cdot p_{i,LC}(a_i \mid f_i, s_i = 0)}.$$

These odds ratios for ever-smokers were taken from the pooled case-control study by Pesch et al.,[6] due to the absence of comparable longitudinal data linking smoking history and lung cancer risk in Singapore.

**Progression in the pre-clinical phase**

Lung cancer stages were defined by the American Joint Committee on Cancer (AJCC) 6th–8th editions depending on the model year. The sojourn time in each stage was independently sampled from a Weibull distribution with a shape parameter of 1.44 and gender- and stage-specific scale parameters adopted from Haaf et al. and further calibrated to histology type distribution from cancer registry data (Table S2).[7] At the end of Stage I, II, or III, individuals either progressed into the next preclinical stage or became symptomatic with a stage-specific probability (Table S3) and got clinically diagnosed, while in Stage IV, only clinical diagnosis was possible.[7] Upon diagnosis, individuals transitioned to the clinical phase, with no further progression between stages thereafter. All individuals who developed lung cancer were assumed to be eventually diagnosed if death from other causes did not occur first.

**Table S2. Gender- and stage-specific scale parameters, along with the corresponding means and standard deviations, of the Weibull distributions for sojourn time.**

| Gender | Stage | Scale parameter | Mean | Standard deviation |
|---|---|---|---|---|
| Male | I | 2.93 | 2.67 | 1.88 |
| | II | 0.55 | 0.50 | 0.35 |
| | III | 0.98 | 0.89 | 0.63 |
| | IV | 0.88 | 0.80 | 0.57 |
| Female | I | 3.28 | 2.98 | 2.06 |
| | II | 0.62 | 0.56 | 0.40 |
| | III | 1.09 | 0.99 | 0.70 |
| | IV | 0.99 | 0.90 | 0.63 |

**Table S3. Stage-specific diagnosis probability for each AJCC lung cancer staging system.**

| Stage | 6th Edition | 7th Edition | 8th Edition |
|---|---|---|---|
| I | 0.87 | 0.91 | 0.86 |
| II | 0.99 | 0.94 | 0.94 |
| III | 0.67 | 0.83 | 0.84 |
| IV | 1.00 | 1.00 | 1.00 |

**Survival in the clinical phase**

A Gompertz proportional hazards model was applied to capture the temporal trends in survival hazards by stage, smoking status, and diagnosis year. Specifically, for individual $i$ with smoking status $s_i$ (0 for never-smoker and 1 for ever-smoker) and diagnosed at stage $k_i$ in calendar year $y_i$, the hazards at time $t$ since diagnosis (calendar year $(y_i + t)$) was modelled as

$$h(t \mid k_i, s_i, y_i) = \exp(\theta_{0,k_i} + \theta_{1,k_i} \cdot t + \theta_2 \cdot 1_{s_i=1} + \theta_3 \cdot y_i),$$

where parameter $\theta_{0,k_i}$ determines the stage-specific scale and $\theta_{1,k_i}$ characterises the stage-specific temporal growth rate.

Let $a_i$ be the age of individual $i$ in calendar year $y_i$, and $m_i(a_i, y_i)$ be the time- and demographic-dependent mortality rate due to other causes, as estimated by DEMOS excluding lung cancer specific deaths. The corresponding hazard was

$$d_i(a_i, y_i) = -\log(1 - m_i(a_i, y_i)).$$

Then the time-dependent total death hazard at time $t$ since diagnosis (calendar year $(y_i + t)$) was calculated as

$$f_i(t \mid a_i, s_i, y_i) = d_i(a_i, y_i + t) + h(t \mid k_i, s_i, y_i).$$

Assuming annual updates of this hazard, the annual death probability was computed as

$$1 - \exp\left(-f(t \mid a_i, s_i, y_i)\right)).$$

Parameters for the natural history model were estimated by optimisation, minimising the weighted sum of squared differences between model-predicted outcomes (incidence, mortality, and survival probabilities) and empirical observations (Figure 2). Baseline age and gender effects in the carcinogenesis model ($\alpha_{0,g}$ and $\gamma_g$) were informed by age- and sex-stratified incidence data from 2009 to 2019, provided by STCC. The survival model parameters ($\theta_{0:1,k}$ and $\theta_{2:3}$) were estimated using individual-level mortality data from STCC for individuals diagnosed between 2008 and 2019, with follow-up through 2023 (Figure S3). Model-projected five-year aggregated incidence and mortality counts were further validated against values reported by NRDO in 2023, with close alignment between projection and observation supporting reliability of the model (Figure S4).

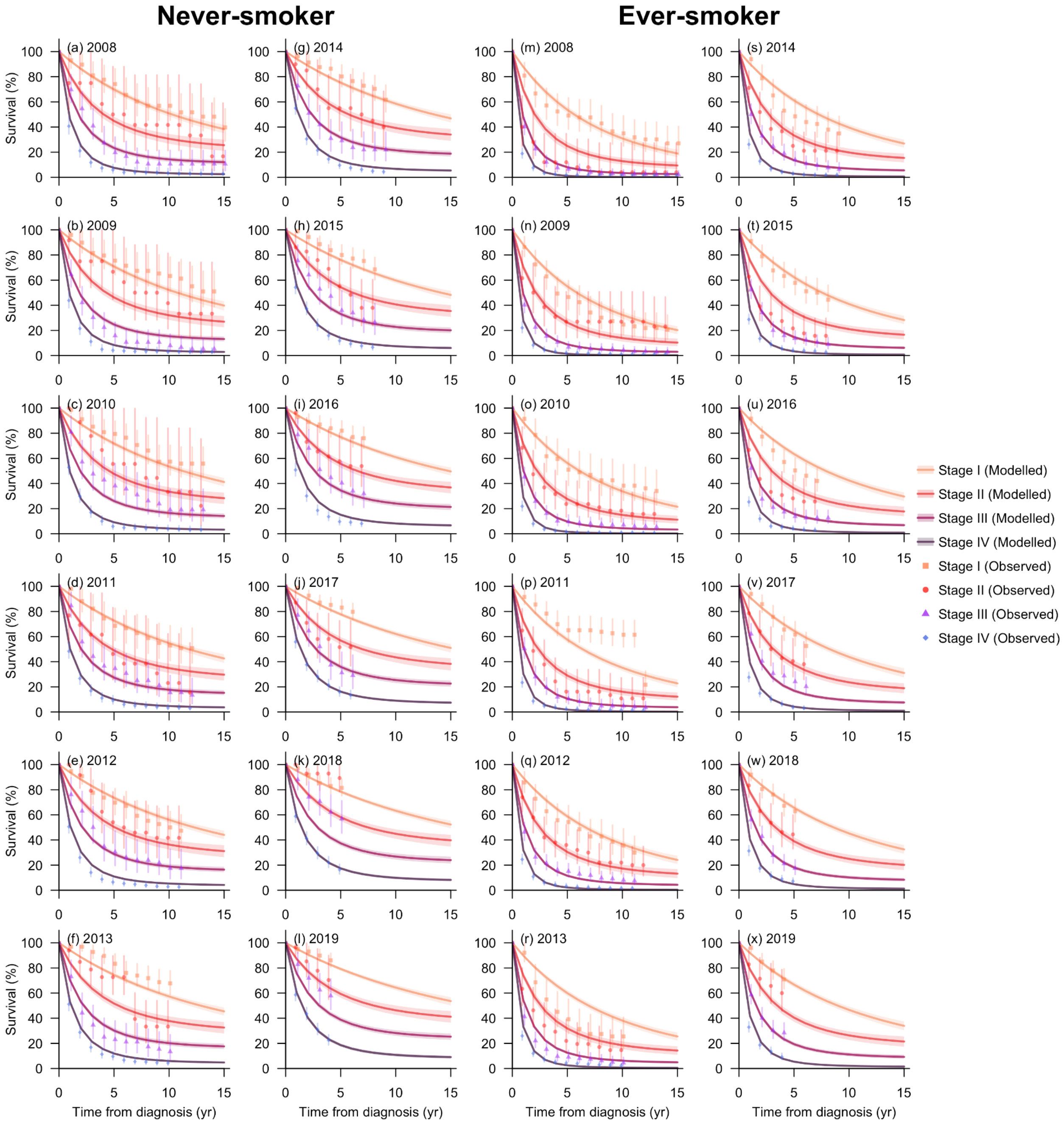

**Figure S3. Lung cancer survival probability by smoking status, stage, and diagnosis year.** The lines represent model-estimated stage-stratified survival probabilities over 15 years of follow-up for cases diagnosed between 2008 and 2019, with shaded areas indicating the corresponding 95% CIs. Observed survival probabilities are shown as points, with vertical lines representing the 95% CIs calculated from reported standard errors.

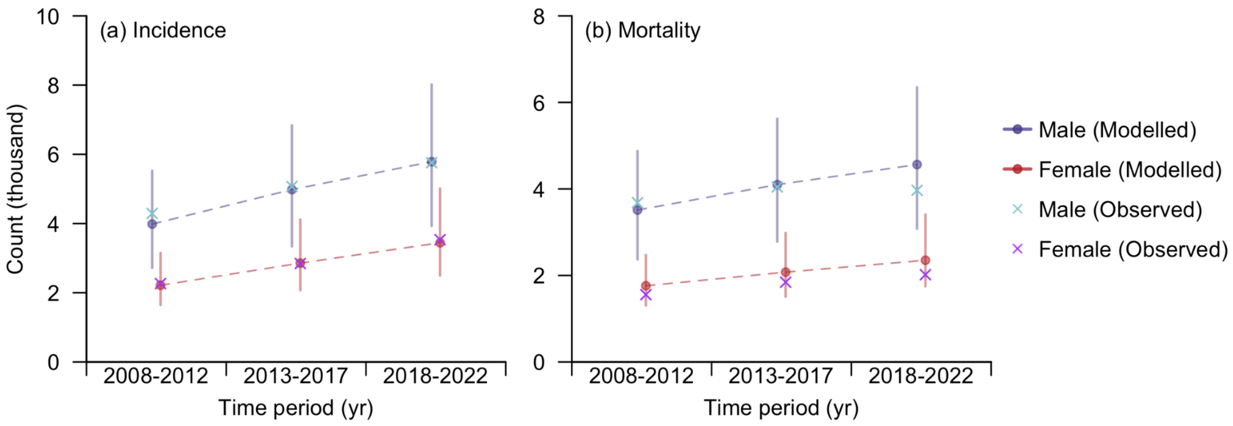

**Figure S4. Lung cancer incidence and mortality over time.** This includes (a) five-year incidence from 2008 to 2022, and (b) five-year deaths from 2008 to 2017. Model-simulated estimates are shown as dots (means) with vertical lines representing uncertainty intervals, while cross marks denote values reported by the National Registry of Diseases Office.

### Health and economic evaluation of screening strategies

#### QALYs

Quality-adjusted life years (QALYs) were calculated as life-years weighted by lung cancer stage, using weights of 1 for healthy individuals, 0.87 for Stages I–II, 0.77 for Stage III, and 0.57 for Stage IV.[8]

#### Late-stage cases

Late-stage cases included all lung cancer diagnoses at Stage III or IV among individuals simulated between 2025 and 2050.

#### Deaths

Deaths were defined as the number of individuals simulated between 2025 and 2050 who died from lung cancer.

#### Overdiagnosis rate

Overdiagnosis rate was calculated as the proportion of screen-detected cases between 2025 and 2050 that would not have become clinically diagnosed (symptomatic) during their lifetime in the absence of screening.

#### False-positive rate

False-positive rate was defined as the proportion of screened individuals without the disease who received a positive screening result among all screenings occurring between 2025 and 2050.

#### Costs

The cost of screening, including the LDCT scan and follow-up consultation, was set at SGD 450 per screen,[10,11] with a possible additional SGD 2,476 incurred by lung biopsy test for false-positive results.[12]

The cost of lung-cancer treatment was modelled as a function of smoking status, diagnosed stage, and time since diagnosis, using a generalized additive model (GAM) fitted to real-world cost data reported by STCC from 2009 to 2019 (Figure S5). The reported costs were converted to 2024 Singapore dollars (SGD) using the Singapore Medical Consumer Price Index (CPI).[13] A generalized additive model (GAM) was employed to fit the data, where the logarithm of the expected treatment cost ($c$) was modelled as a function of smoking status ($s$), diagnosed lung cancer stage ($k$), and time since diagnosis ($t$):

$$\log c = \phi_0 + \phi_1 \cdot 1_{s \neq never\ smoker} + \phi_2 \cdot k + \psi(t).$$

In this, $\psi(\cdot)$ represents a spline-smoothed temporal effect, capturing the non-linear trend in treatment costs over time following diagnosis. Model parameters were estimated using the restricted maximum likelihood (REML) algorithm, with stratum-specific weights (defined by smoking status, cancer stage, and time since diagnosis) applied to adjust for unequal sample sizes across subgroups. Uncertainty in parameter estimates was addressed through bootstrapping the observed costs, generating 100 parameter draws for subsequent cost calculations.

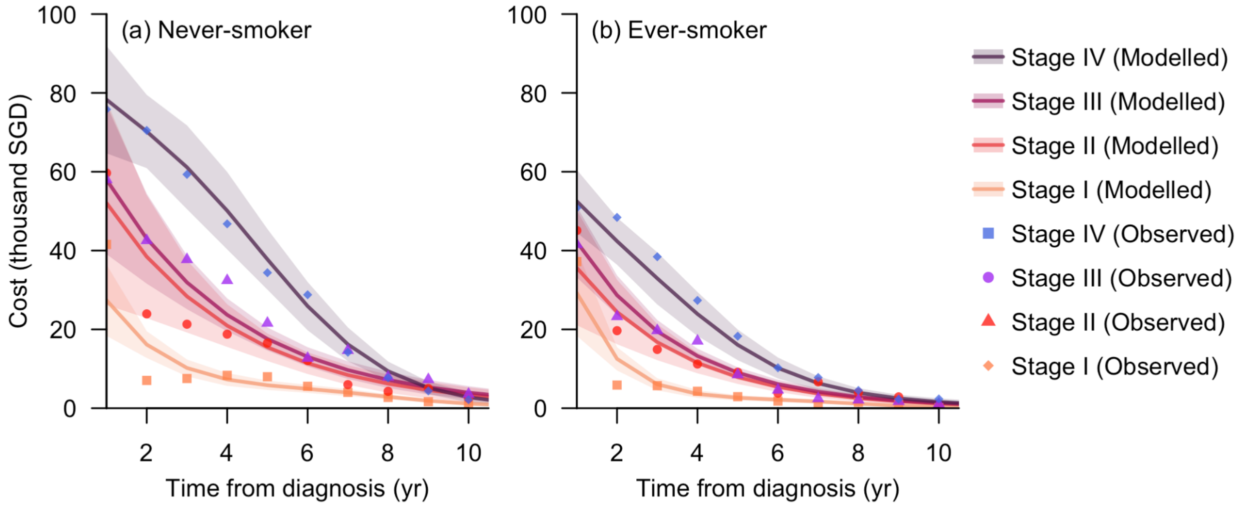

**Figure S5. Comparison of model-predicted and observed treatment costs, stratified by smoking status, diagnosed stage, and time from diagnosis.** Lines represent the predicted means, and shaded areas indicate the corresponding 95% CIs from the fitted model.

**Table S4. Parameters of the screening model.**

| Parameter | Value | Source |
|---|---|---|
| Sensitivity | Stage I: 0.40<br>Stage II: 0.44<br>Stage III: 0.75<br>Stage IV: 0.98 | Ten Haaf et al.[7] |
| Specificity | 0.73 | Kowada et al.[8] |
| Minimum eligible age (years) | 50, 55, 60 | Assumed |
| Maximum eligible age (years) | 75, 80, 85 | Assumed |
| Uptake | 40%, 70%, 100% | Assumed |
| Pack-year (for current smokers) | 20 | US Preventive Services Task Force[9] |
| Quitting duration (years, for former smokers) | 15 | US Preventive Services Task Force[9] |
| Frequency | Annual, biennial, triennial, quadrennial, quinquennial | Assumed |

**Table S5. Parameters of the economic model.**

| Parameter | Value | Source |
|---|---|---|
| Health utility | Stage I: 0.87<br>Stage II: 0.87<br>Stage III: 0.77<br>Stage IV: 0.57<br>Death: 0 | Kowada et al.[8] |
| LDCT scan cost (SGD) | 450 | ATA Medical and Health365.SG[10,11] |
| Lung Biopsy cost (SGD) | 2476 | Ministry of Health[12] |

| | | |
|---|---|---|
| Annual discounting rate | 3% | Chootipongchaivat et al.[14] |

**Evaluation metrics and scores**

The screening strategies were assessed using six metrics: QALYs, late-stage incidence averted, deaths averted, additional costs, overdiagnosis rate, and false-positive rate.

For each combination of gender and tobacco control scenario, four screening strategies were selected from the 135 candidates (defined by 3 starting ages, 3 cessation ages, 3 uptake rates, and 5 screening frequencies) based on the following decision criteria:

1) Most life-saving with ≤10% overdiagnosis rate (MLSOD10): the strategy that achieved the highest QALY gains relative to the baseline, among those with an overdiagnosis rate not exceeding 10%;
2) Most cost-effective: the strategy with the lowest ICER;
3) Lowest-overdiagnosis: the strategy with the lowest overdiagnosis rate;
4) Overall optimal (TOPSIS-recommended; TRS): the strategy identified using TOPSIS (Technique for Order Preference by Similarity to Ideal Solution) algorithm among all Pareto-efficient candidates.

The first three strategies were selected using paired t-tests, where strategies that were statistically significantly superior (higher or lower, as appropriate) to the greatest number of other strategies were identified. When multiple strategies met this criterion, the strategy with the highest mean score was selected. For the fourth strategy, the selection process is as below:[15]

1) Scores for each metric were calculated by log-transforming the values (to mitigate the influence of extreme values) and rescaling to a 0–1 range across all screening strategies within each combination of gender and tobacco control scenario using min-max normalisation (i.e., $(x - min)/(max - min)$). For metrics where larger values indicated less desirable outcomes (additional costs, overdiagnosis rate, and false-positive rate), scores were inverted as $(1 - scaled\ value)$, such that higher scores consistently reflect more favourable performance.
2) Pareto efficient screening strategies were selected as those with an overdiagnosis rate not significantly higher than 10% (i.e., the estimated upper bound for the mean rate across the 100 simulations was below 10%) and that were not dominated across all six scores (i.e., for each strategy, no other strategy achieved statistically significantly higher scores for all six metrics).
3) Within each round of simulations, the TOPSIS scores for individual screening strategies were calculated in five steps:
    a. Let $x_{ij}$ denote the score of metric $j$ for strategy $i$, the normalised matrix $R = (r_{ij})$ was constructed as $r_{ij} = x_{ij}/\sqrt{\sum_i x_{ij}^2}$ ;
    b. Equal weights were assigned to all metrics. The ideal and negative ideal solutions for each metric were derived: $r_j^+ = \max_i(r_{ij})$, $r_j^- = \min_i(r_{ij})$;
    c. The Euclidean distances of each candidate to the ideal and negative ideal solutions were computed as $S_i^+ = \sqrt{\sum_j (r_{ij} - r_j^+)^2}$ and $S_i^- = \sqrt{\sum_j (r_{ij} - r_j^-)^2}$;
    d. The closeness coefficient was calculated as $C_i = S_i^+/(S_i^+ + S_i^-)$.
4) The overall TOPSIS score for each strategy was computed as the mean closeness coefficient across the 100 simulations. The strategy with the highest TOPSIS score was selected as optimal.

This TOPSIS-ranked strategy represented the best overall balance across all six metrics, achieving health gains while controlling costs, overdiagnosis, and false positives.

### Main analysis results

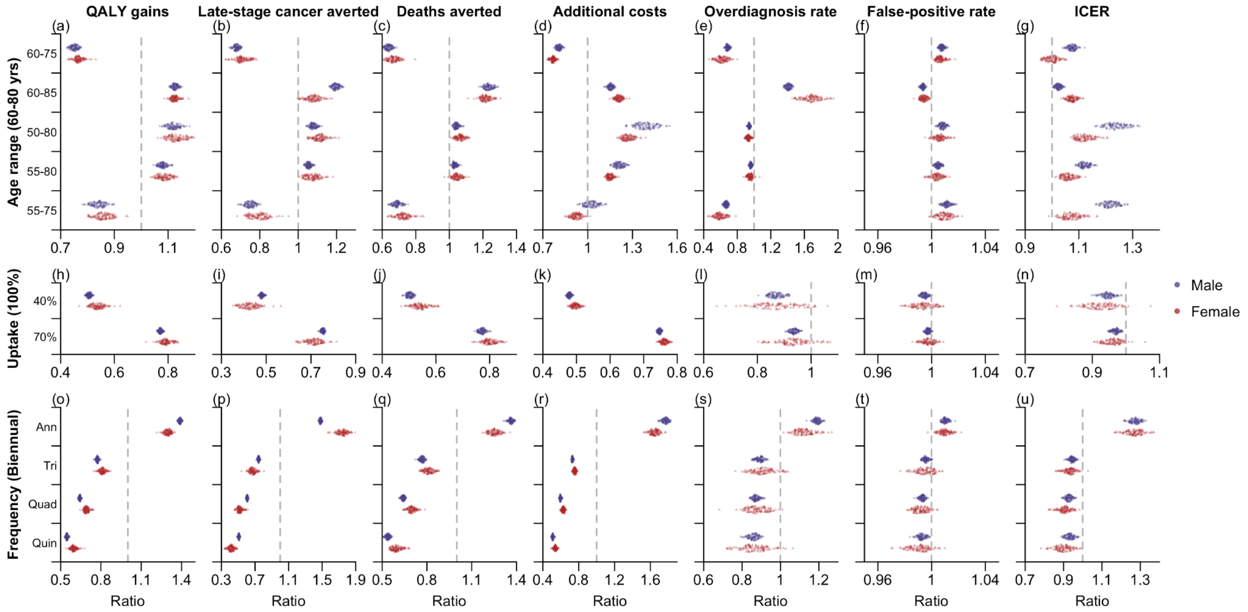

**Figure S6. Comparison of TRS with alternative screening strategies.** Ratios of outcomes for each alternative strategy relative to TRS (denominator; biennial screening of eligible ever-smokers aged 60–80 years) were computed per simulation and shown as dots, with blue and red indicating males and females, respectively. Each alternative strategy differs from TRS in one attribute (age range, uptake, or screening frequency), with reference values given in parentheses on the left and alternative values listed on the y-axis. All strategies were evaluated over 2025–2050 across seven metrics (Columns 1–7): QALY gains, late-stage cancers averted, deaths averted, additional costs, overdiagnosis rate, false-positive rate, and ICER, each relative to the scenario without additional screening.

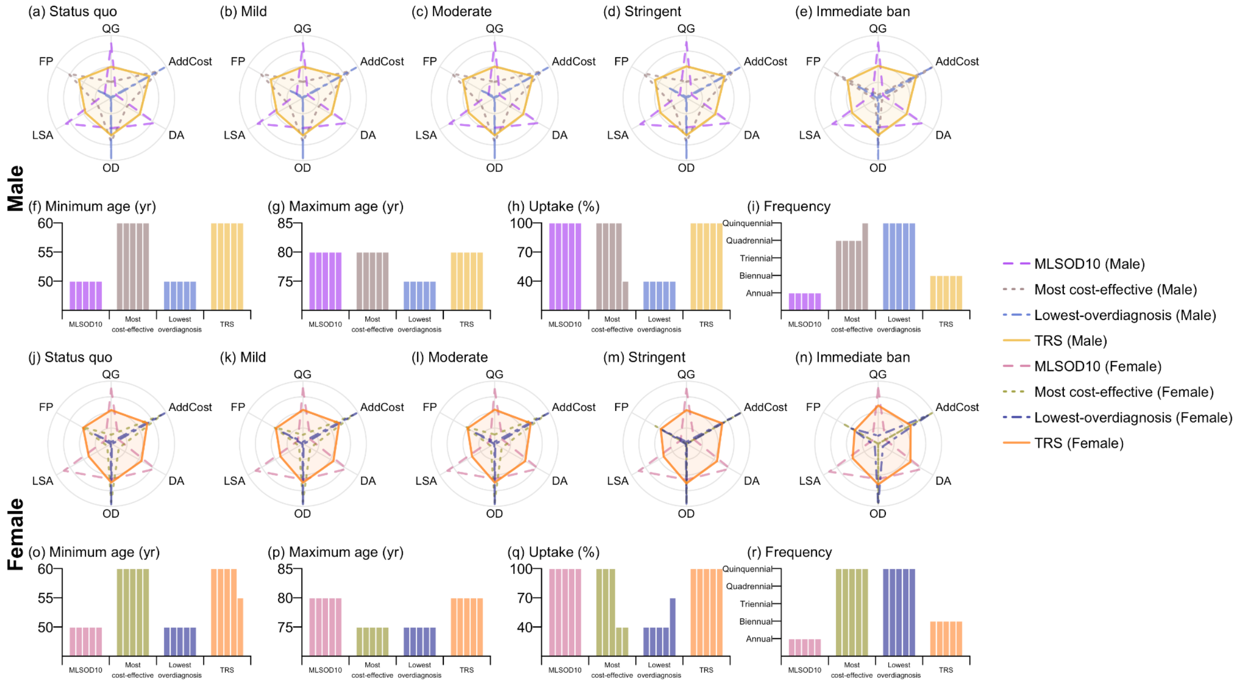

**Figure S7. Recommended screening strategies under four decision criteria.** The four selected strategies comprise the most life-saving with 10% overdiagnosis rate (MLSOD10), the most cost-effective, the lowest-overdiagnosis, and the overall optimal strategy (TRS). Subfigures (a)–(e) (Row 1, for male ever-smokers) and (j)–(n) (Row 3, for female ever-smokers) present the scores for six metrics evaluated over year 2025–2050: QALY gains (QG), late-stage cancer averted (LSA), deaths averted (DA), and additional costs (AddCost) relative to the scenario without additional screening, as well as overdiagnosis rate (OD) and false positive rate (FP). All the metrics represent medians across simulations and are rescaled to a 0–1 range. Subfigures (f)–(i) (Row 2, for male ever-smokers) and (o)–(r) (Row 4, for female ever-smokers) show the characteristics of the selected strategies, including minimum age, maximum age, screening uptake rate, and screening frequency (rescaled to an annual rate). The five adjacent columns within each decision criteria correspond to the five tobacco control scenarios ranging from status quo to immediate ban. Strategies selected under different criteria for male and female ever-smokers are distinguished by colour and line style, as indicated in the legend on the right.

**Table S6. Lung cancer burden and associated total costs across five tobacco control scenarios without additional screening.** Variables to quantify lung cancer burden include QALYs, number of late-stage cases, and deaths. All values are reported as medians and 95% uncertainty intervals summarised from the 100 simulations.

| **Scenario** | **QALYs (000s)** | **Late-stage cases (000s)** | **Deaths (000s)** | **Total costs (billion SGD)** |
|---|---|---|---|---|
| Male | | | | |
| Status quo | 85.73 (85.48–85.88) | 134.72 (102.15–191.09) | 68.43 (51.94–90.89) | 10.92 (8.27–14.99) |
| Mild | 85.74 (85.5–85.89) | 130.73 (99.42–184.27) | 66.51 (50.42–88.92) | 10.73 (8.15–14.78) |
| Moderate | 85.75 (85.52–85.90) | 124.9 (95.04–174.56) | 63.53 (48.21–85.47) | 10.44 (7.94–14.45) |
| Stringent | 85.78 (85.55–85.92) | 118.86 (90.55–164.61) | 59.67 (45.80–80.99) | 10.05 (7.66–13.97) |
| Immediate ban | 85.81 (85.59–85.94) | 113.45 (86.31–155.03) | 55.05 (42.65–74.76) | 9.59 (7.29–13.34) |
| Female | | | | |
| Status quo | 85.71 (85.46–85.86) | 138.88 (106.61–198.19) | 71.06 (53.67–92.2) | 10.33 (7.75–14.16) |
| Mild | 85.72 (85.48–85.87) | 135.35 (103.70–190.33) | 69.1 (52.41–90.18) | 10.15 (7.64–13.96) |
| Moderate | 85.74 (85.51–85.88) | 129.45 (98.13–179.41) | 65.56 (50.27–86.88) | 9.86 (7.46–13.64) |
| Stringent | 85.76 (85.54–85.90) | 122.56 (92.75–168.37) | 61.19 (47.35–82.26) | 9.51 (7.21–13.20) |
| Immediate ban | 85.80 (85.58–85.93) | 115.82 (87.75–157.42) | 56.26 (43.67–75.71) | 9.11 (6.86–12.67) |

**Table S7. Values (medians and 95% UIs) of seven evaluation metrics for MLSOD10 across five tobacco control scenarios.** These metrics include QALY gains (QG), late-stage cancer averted (LSA), deaths averted (DA), additional costs (AddCost), overdiagnosis rate (OD), false positive rate (FP), and ICER. They were calculated with reference to the setting without additional screening with each tobacco control scenario.

| Scenario | QG (000s) | LSA (000s) | DA (000s) | AddCost (million SGD) | OD (%) | FP (%) | ICER (000s SGD/QALY) |
|---|---|---|---|---|---|---|---|
| Male | | | | | | | |
| Status quo | 45.94 (31.55–66.01) | 11.11 (7.59–15.91) | 5.25 (3.60–7.61) | 2.03 (1.72–2.39) | 9.67 (9.14–10.39) | 26.15 (26.00–26.29) | 44.14 (32.25–60.15) |
| Mild | 44.65 (30.33–64.2) | 10.67 (7.21–15.33) | 5.04 (3.44–7.31) | 1.99 (1.7–2.36) | 9.64 (9.09–10.31) | 26.16 (26.00–26.29) | 44.95 (32.81–61.21) |
| Moderate | 42.27 (28.11–60.61) | 9.77 (6.54–14.12) | 4.60 (3.14–6.77) | 1.93 (1.64–2.3) | 9.57 (9.03–10.21) | 26.17 (26.02–26.31) | 46.49 (34.09–63.40) |
| Stringent | 36.82 (24.32–53.41) | 8.19 (5.35–11.95) | 3.82 (2.60–5.75) | 1.79 (1.52–2.17) | 9.37 (8.68–9.97) | 26.19 (26.05–26.33) | 49.73 (36.46–67.30) |
| Immediate ban | 26.35 (18.03–38.76) | 5.49 (3.65–7.89) | 2.59 (1.77–3.87) | 1.49 (1.26–1.79) | 8.81 (8.28–9.31) | 26.24 (26.12–26.35) | 56.67 (41.12–74.64) |
| Female | | | | | | | |
| Status quo | 5.14 (2.48–10.52) | 0.85 (0.40–1.79) | 0.63 (0.28–1.26) | 0.22 (0.13–0.38) | 7.71 (6.34–8.73) | 26.15 (25.95–26.35) | 43.00 (29.76–57.99) |
| Mild | 4.93 (2.30–10.10) | 0.80 (0.36–1.70) | 0.59 (0.26–1.20) | 0.21 (0.12–0.37) | 7.63 (6.34–8.72) | 26.16 (25.95–26.36) | 44.00 (30.39–58.85) |
| Moderate | 4.40 (2.02–9.31) | 0.70 (0.31–1.51) | 0.52 (0.23–1.09) | 0.20 (0.11–0.35) | 7.63 (6.32–8.68) | 26.18 (25.96–26.39) | 45.89 (31.28–60.68) |
| Stringent | 3.72 (1.70–7.85) | 0.55 (0.25–1.20) | 0.41 (0.18–0.87) | 0.18 (0.10–0.32) | 7.43 (5.88–8.46) | 26.20 (25.97–26.41) | 48.21 (33.05–63.23) |
| Immediate ban | 2.70 (1.37–5.34) | 0.36 (0.18–0.70) | 0.28 (0.14–0.54) | 0.14 (0.08–0.24) | 7.01 (5.41–8.17) | 26.22 (25.98–26.45) | 51.49 (35.33–69.75) |

**Table S8. Values (medians and 95% UIs) of seven evaluation metrics for TRS across five tobacco control scenarios.** These metrics include QALY gains (QG), late-stage cancer averted (LSA), deaths averted (DA), additional costs (AddCost), overdiagnosis rate (OD), false positive rate (FP), and ICER. They were calculated with reference to the setting without additional screening with each tobacco control scenario.

| Scenario | QG (000s) | LSA (000s) | DA (000s) | AddCost (million SGD) | OD (%) | FP (%) | ICER (000s SGD/QALY) |
|---|---|---|---|---|---|---|---|
| Male | | | | | | | |
| Status quo | 29.33 (19.95–42.76) | 6.96 (4.64–9.95) | 3.67 (2.47–5.35) | 0.81 (0.65–1.01) | 8.51 (8.06–9.10) | 25.73 (25.44–25.98) | 26.96 (20.01–35.97) |
| Mild | 28.43 (19.2–41.62) | 6.68 (4.41–9.57) | 3.51 (2.37–5.17) | 0.80 (0.64–1.00) | 8.46 (8.00–9.05) | 25.74 (25.46–25.99) | 27.40 (20.40–36.44) |
| Moderate | 26.77 (17.83–39.18) | 6.13 (4.00–8.82) | 3.22 (2.16–4.81) | 0.78 (0.62–0.98) | 8.37 (7.93–9.00) | 25.77 (25.49–26.02) | 28.51 (21.34–37.88) |
| Stringent | 23.42 (15.32–34.56) | 5.10 (3.30–7.49) | 2.70 (1.79–4.10) | 0.73 (0.58–0.92) | 8.20 (7.67–8.78) | 25.83 (25.56–26.07) | 30.59 (23.06–40.57) |
| Immediate ban | 17.27 (11.51–25.15) | 3.49 (2.26–4.99) | 1.81 (1.20–2.71) | 0.61 (0.49–0.76) | 7.75 (7.18–8.26) | 25.92 (25.69–26.14) | 34.98 (26.22–45.55) |
| Female | | | | | | | |
| Status quo | 3.51 (1.65–7.28) | 0.45 (0.20–0.94) | 0.46 (0.21–0.95) | 0.10 (0.06–0.19) | 7.16 (5.73–8.46) | 25.80 (25.34–26.15) | 29.70 (21.39–38.54) |
| Mild | 3.37 (1.54–7.00) | 0.42 (0.19–0.89) | 0.44 (0.19–0.92) | 0.10 (0.05–0.18) | 7.11 (5.73–8.57) | 25.83 (25.32–26.18) | 30.36 (21.88–39.06) |
| Moderate | 3.07 (1.35–6.51) | 0.37 (0.16–0.80) | 0.39 (0.17–0.84) | 0.09 (0.05–0.18) | 7.02 (5.75–8.49) | 25.84 (25.38–26.21) | 31.40 (22.55–40.07) |
| Stringent | 2.56 (1.14–5.47) | 0.29 (0.12–0.63) | 0.31 (0.14–0.67) | 0.08 (0.05–0.16) | 6.85 (5.49–8.18) | 25.90 (25.43–26.25) | 32.84 (23.90–42.33) |
| Immediate ban | 1.89 (0.93–3.74) | 0.19 (0.09–0.37) | 0.21 (0.11–0.42) | 0.07 (0.04–0.12) | 6.43 (5.07–7.98) | 25.93 (25.46–26.39) | 35.37 (25.68–46.15) |

**Sensitivity analysis: Stage-specific survival rate extrapolation**

In the main analysis, we extrapolated stage-specific survival hazards from observations between 2009 and 2019, modelling them to decrease exponentially with increasing calendar year at diagnosis. This approach assumes that trends in technological advances and improvements in treatment effectiveness remain time-invariant beyond the observation period. To examine how this assumption affected projected mortality and screening impacts, in this sensitivity analysis we conservatively assumed that the stage-specific survival hazards for cases diagnosed between 2025 and 2050 remained the same as 2024 levels. Specifically, for individual $i$ with smoking status $s_i$ and diagnosed at stage $k_i$ in calendar year $y_i$, the hazards at time $t$ since diagnosis (calendar year $(y_i + t)$) was calculated as

$$h(t|k_i, s_i, y_i) = \exp(\theta_{0,k_i} + \theta_{1,k_i} \cdot t + \theta_2 \cdot 1_{s_i=1} + \theta_3 \cdot \min(y_i, 2024)).$$

The projected results suggested a mild increase in incidence and mortality over the 26-year period, along with smaller QALY gains and lower costs associated with screening and treatment (Figure S8–S9). However, the association between screening intensity and health impacts, as well as the resulting screening strategy recommendations, remained largely unchanged (Figure S9–S10).

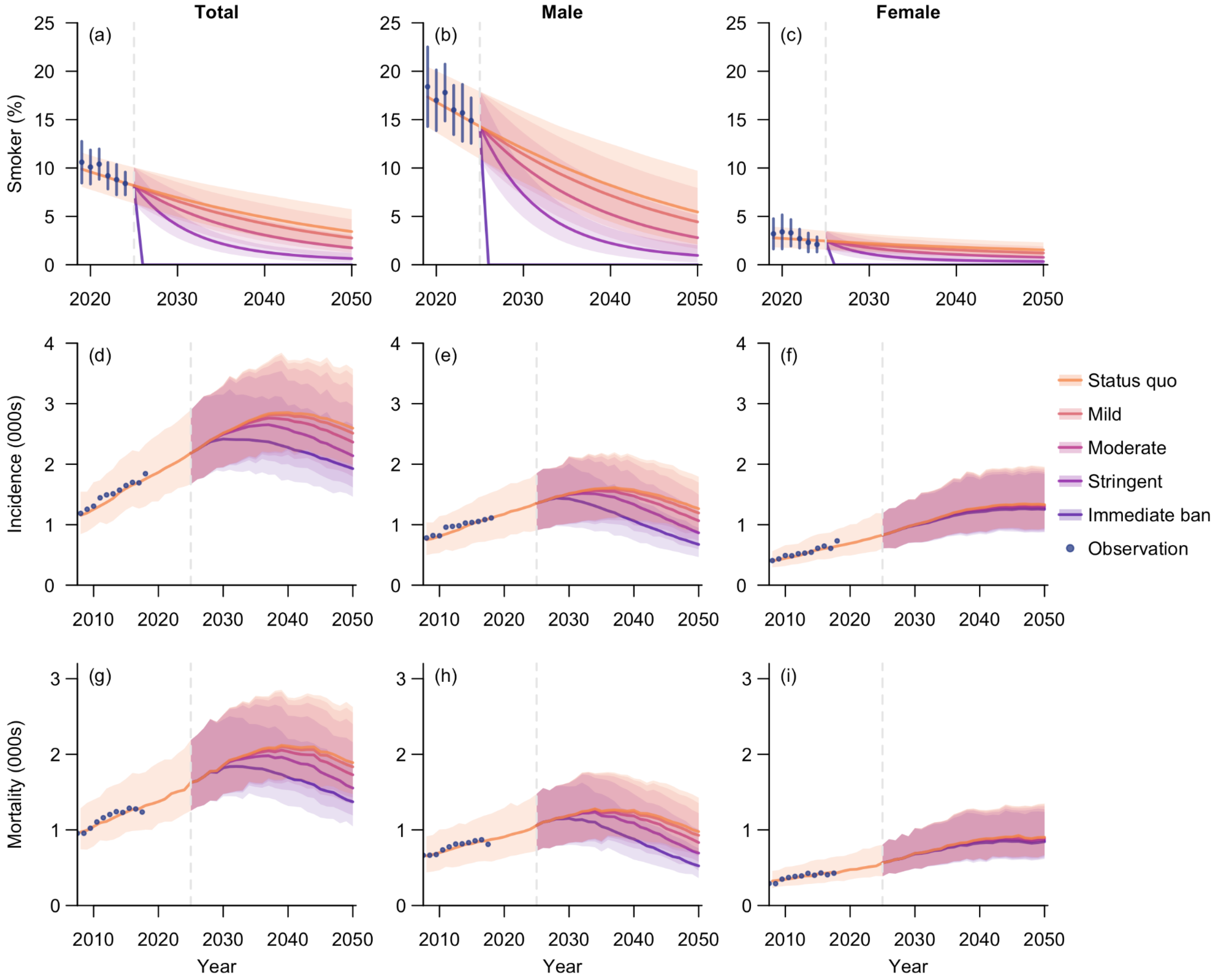

**Figure S8. Baseline projections under five tobacco control scenarios when conservatively assuming that survival rates remained at 2024 levels.** Row 1 shows projected smoking prevalence from 2019 to 2050, while Rows 2 and 3 lung show cancer incidence and mortality, respectively, from 2008 to 2050. Results are presented for the total population (Column 1) and stratified by gender (Columns 2–3). Tobacco control campaigns were assumed to initiate in 2025, without screening implemented in any scenario. Projected medians are represented by lines, with shades indicating the corresponding 95% uncertainty intervals. Note the different time scales used in Row 1 and Row 2–3.

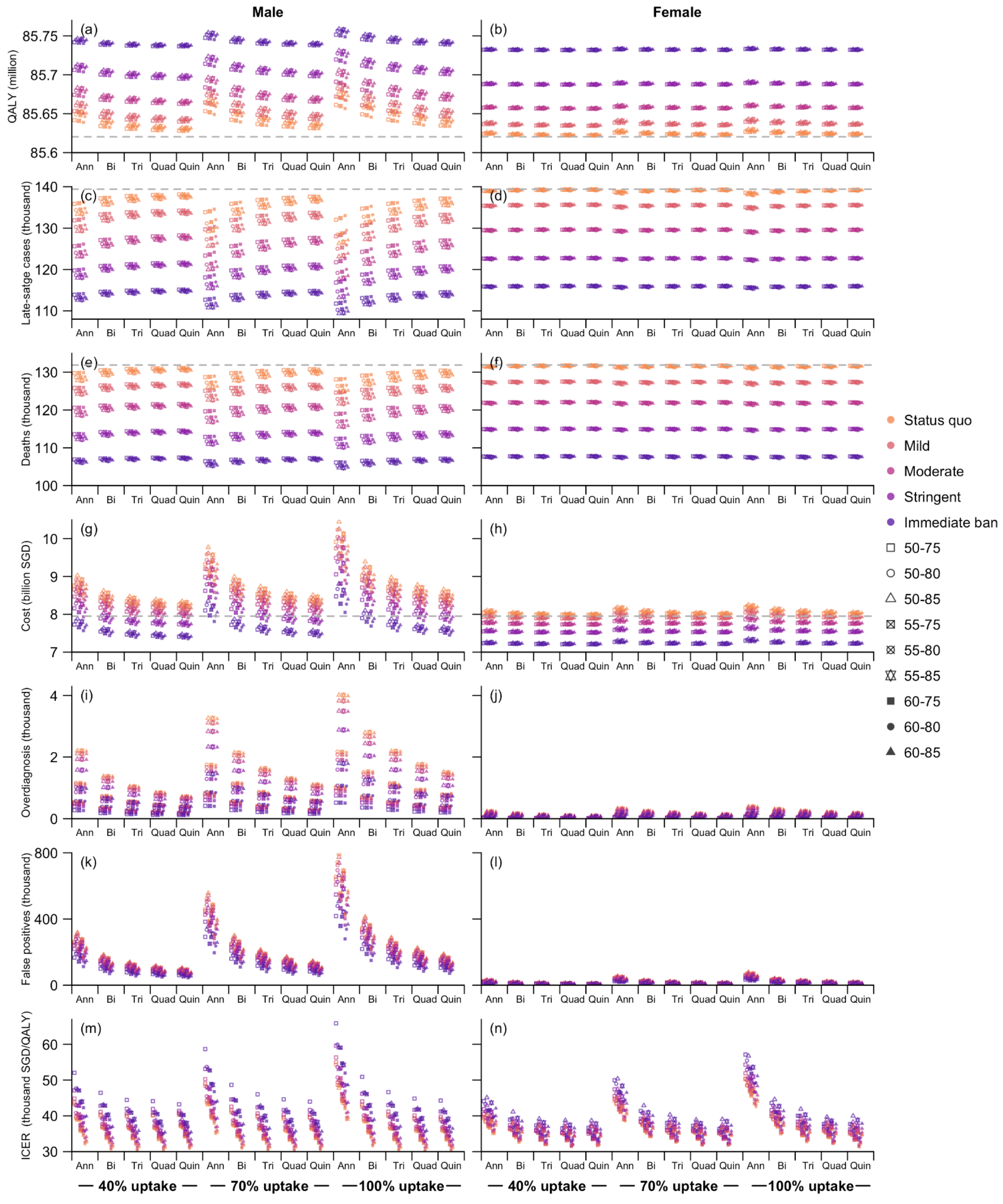

**Figure S9. Comparison of screening strategies across five tobacco control scenarios when conservatively assuming that survival rates remained at 2024 levels.** All strategies were evaluated over 2025–2050 using seven metrics (Rows 1–7): total QALYs, late-stage cancers, deaths, costs, overdiagnosed cases, false-positives cases, and ICER. Points show median estimates across simulations for each strategy. Strategies are defined by target age range, screening frequencies, and uptake, with frequency-uptake combinations shown on the x-axis and age range indicated by different point styles (see legend). The grey vertical lines in subfigures in subfigures (a)–(h) (Rows 1–4) denote the corresponding values under the status quo scenario without screening. Note that ICERs in subfigure (m)–(n) were for screening only, calculated relative to no additional screening within each tobacco control setting.

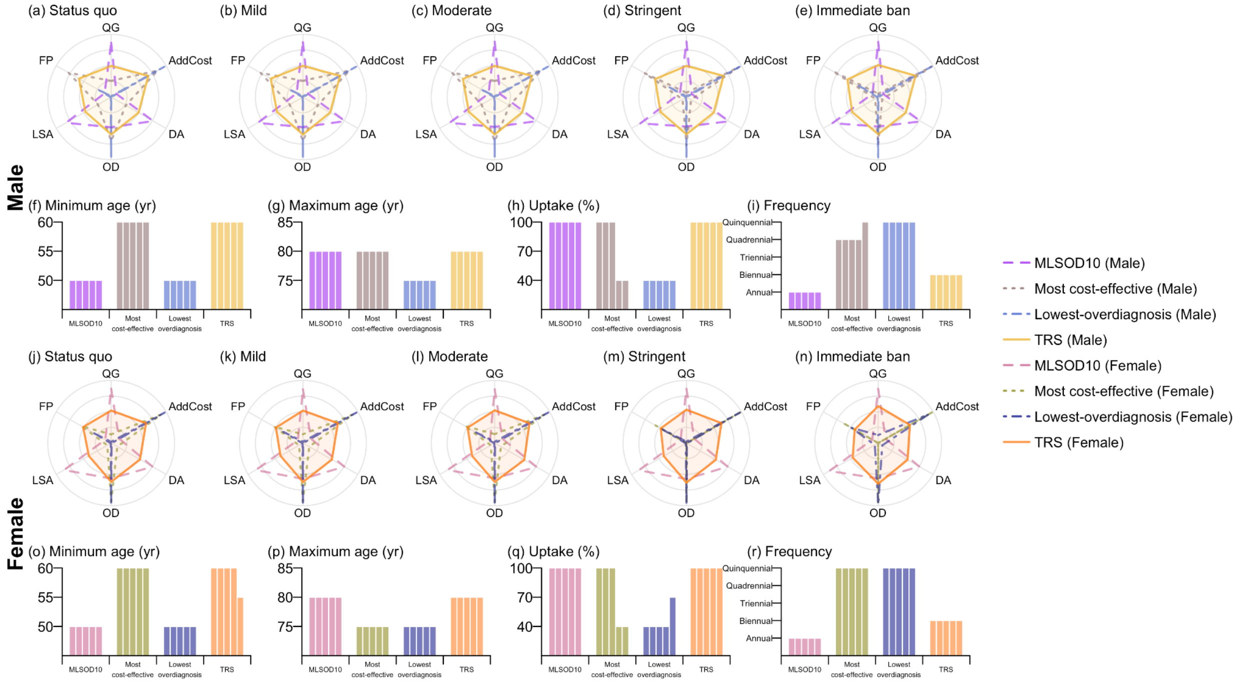

**Figure S10. Recommended screening strategies under four decision criteria when conservatively assuming that survival rates remained at 2024 levels.** The four selected strategies comprise the most life-saving with 10% overdiagnosis rate (MLSOD10), the most cost-effective, the lowest-overdiagnosis, and the overall optimal strategy (TRS). For male ever-smokers, subfigures (a)–(e) (Row 1, for male ever-smokers) and (j)–(n) (Row 3, for female ever-smokers) present the scores for six metrics evaluated over year 2025–2050: QALY gains (QG), late-stage cancer averted (LSA), deaths averted (DA), and additional costs (AddCost) relative to the scenario without additional screening, as well as overdiagnosis rate (OD) and false positive rate (FR). All the metrics represent medians across simulations and are rescaled to a 0–1 range. Subfigures (f)–(i) (Row 2, for male ever-smokers) and (o)–(r) (Row 4, for female ever-smokers) show the characteristics of the selected strategies, including minimum age, maximum age, screening uptake rate, and screening frequency (rescaled to an annual rate). The five adjacent columns within each decision criteria correspond to the five tobacco control scenarios ranging from status quo to immediate ban. Strategies selected under different criteria for male and female ever-smokers are distinguished by colour and line style, as indicated in the legend on the right.

**Sensitivity analysis: Screening sensitivity**

Owing to the high uncertainty in screening sensitivity for Stage I and II disease, we conducted this sensitivity analysis to evaluate how varying sensitivity assumptions affect projected screening benefits. We examined two alternative sets of stage-specific screening sensitivity levels, one lower and the other higher than those assumed in the main analysis (Table S9).[7]

**Table S9. Screening sensitivity by stage.**

| Stage | Main analysis | Low-sensitivity scenario | High-sensitivity scenario |
|---|---|---|---|
| I | 0.40 | 0.38 | 0.44 |
| II | 0.44 | 0.41 | 0.48 |
| III | 0.75 | 0.72 | 0.79 |
| IV | 0.98 | 0.96 | 1.00 |

The projected outcomes suggested improved cost-effectiveness with higher screening sensitivity (Figure S11). Nevertheless, variations in screening sensitivity had a limited impact on the relative ranking of the screening strategies and the score-based recommendations (Figure S11–S12).

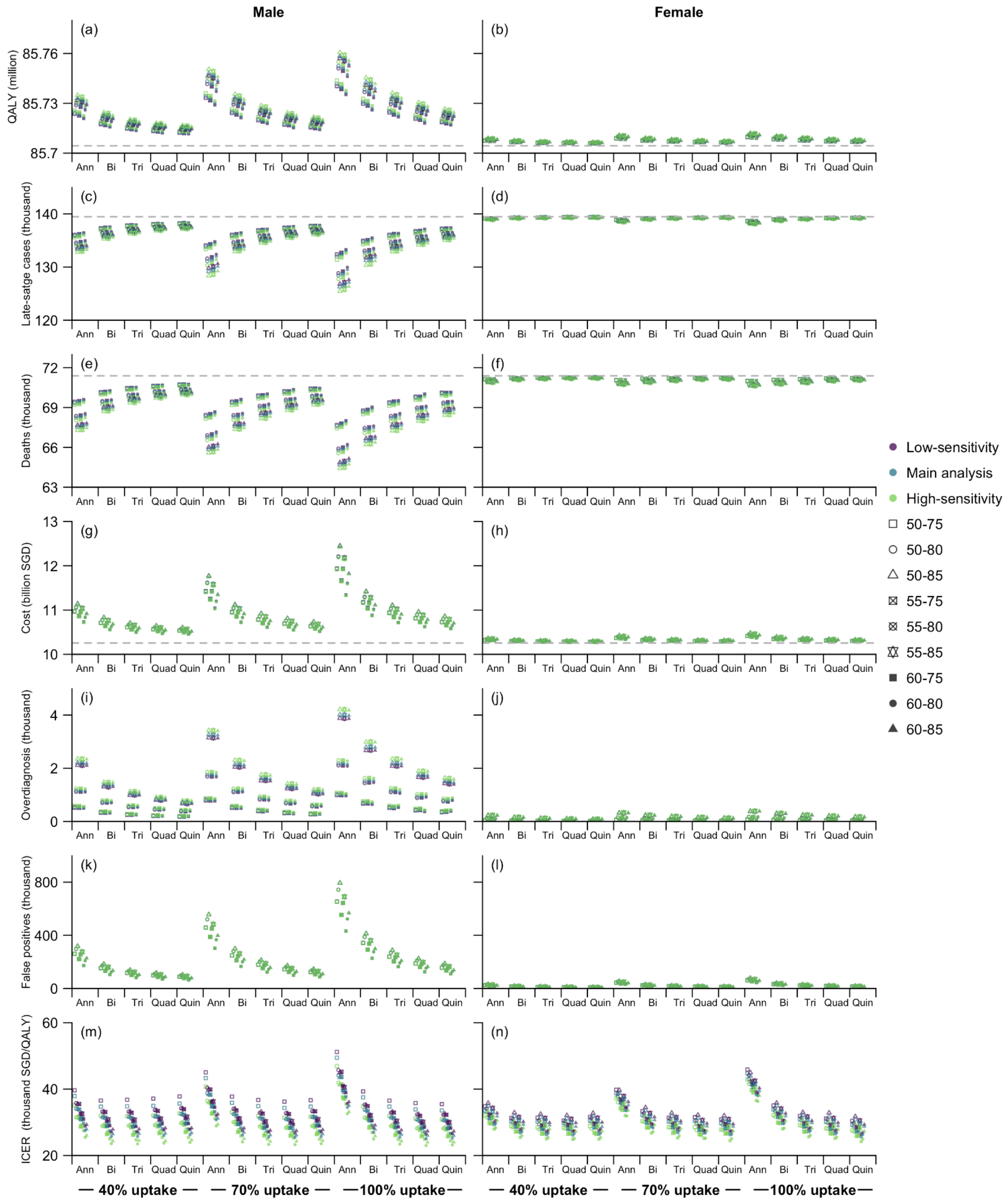

**Figure S11. Comparison of screening strategies across five tobacco control scenarios, assuming different screening sensitivity levels.** All strategies were evaluated over 2025–2050 using seven metrics (Rows 1–7): total QALYs, late-stage cancers, deaths, costs, overdiagnosed cases, false-positives cases, and ICER. Points show median estimates across simulations for each strategy. Strategies are defined by target age range, screening frequencies, and uptake, with frequency-uptake combinations shown on the x-axis and age range indicated by different point styles (see legend). The grey vertical lines in subfigures in subfigures (a)–(h) (Rows 1–4) denote the corresponding values under the status quo scenario without screening. Note that ICERs in subfigure (m)–(n) were for screening only, calculated relative to no additional screening within each tobacco control setting.

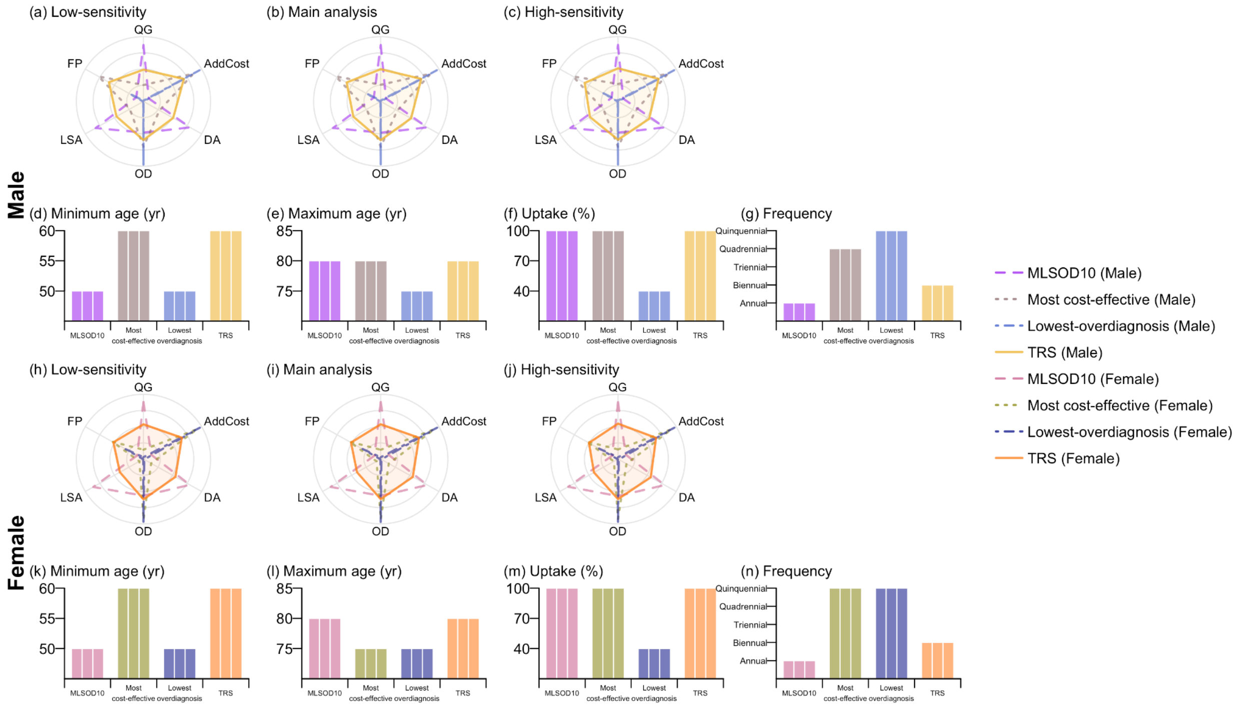

**Figure S12. Recommended screening strategies under four decision criteria for the status quo scenario, assuming different screening sensitivity levels.** The four selected strategies comprise the most life-saving with 10% overdiagnosis rate (MLSOD10), the most cost-effective, the lowest-overdiagnosis, and the overall optimal strategy (TRS). Subfigures (a)–(c) (Row 1, for male ever-smokers) and (h)–(j) (Row 3, for female ever-smokers) present the scores for six metrics evaluated over year 2025–2050: QALY gains (QG), late-stage cancer averted (LSA), deaths averted (DA), and additional costs (AddCost) relative to the scenario without additional screening, as well as overdiagnosis rate (OD) and false positive rate (FP). All the metrics represent medians across simulations and are rescaled to a 0–1 range. Subfigures (d)–(g) (Row 2, for male ever-smokers) and (k)–(n) (Row 4, for female ever-smokers) show the characteristics of the selected strategies, including minimum age, maximum age, screening uptake rate, and screening frequency (rescaled to an annual rate). The three adjacent columns within each decision criteria correspond to the three screening sensitivity levels. Strategies selected under different criteria for male and female ever-smokers are distinguished by colour and line style, as indicated in the legend on the right.

**Sensitivity analysis: eligibility for screening ever-smokers**

In the main analysis, screening eligibility for ever-smokers was defined as current smokers with at least 20 pack-years of exposure or former smokers who had quit smoking within the past 15 years. With the decreasing smoking prevalence over time, it might be feasible to relax these thresholds to enable a broader group of at-risk individuals to benefit from screening.

Accordingly, we performed this sensitivity analysis to evaluate the health and economic impacts resulting from varying the pack-year and quitting-duration thresholds. Specifically, we fixed the screening strategy as either MLSOD10 (annual screening of all ever-smokers aged 50–80 years) or TRS (biennial screening of all ever-smokers aged 60–80 years). We performed microsimulations across pack-year thresholds ranging from 1 to 35 and quitting duration thresholds of 15 and 20 years under all five tobacco control scenarios considered in the main analysis.

For both strategies, the projections suggested a general decline in overall health and economic benefits as the pack-year and quitting-duration thresholds increased, accompanied by improved cost-effectiveness. Screening remained cost-effective, with the mean ICERs across 100 simulations not exceeding the willingness-to-pay threshold of 0.12 million SGD/QALY for all eligibility criteria (Figure S13–S14).

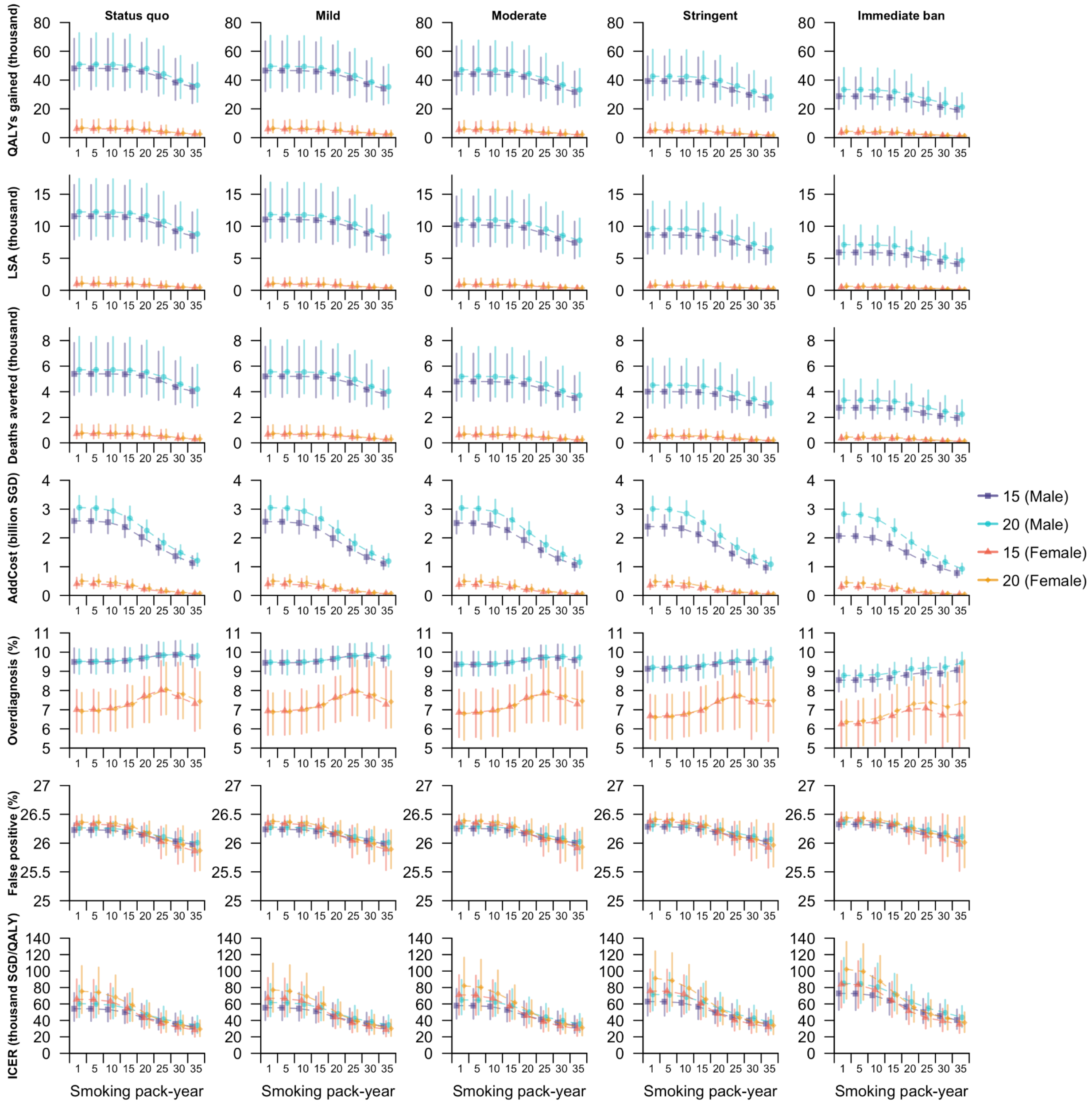

**Figure S13. Projected health and economic impacts of MLSOD10 when varying screening eligibility criteria for ever-smokers.** Outcomes of the screening strategies targeting all ever-smokers aged 50–80 years with annual screening frequency were evaluated over 2025–2050 using seven metrics (one per row), including QALY gains, late-stage cancers averted (LSA), deaths averted, additional costs (AddCost), overdiagnosis rate, false positive rate, and ICER. All these metrics were calculated related to the no-additional-screening scenario. For each metric, dots represent the median across simulations, and vertical lines indicate the corresponding 95% uncertainty intervals (UI). Columns correspond to the five tobacco control scenarios. For current smokers, eight pack-year thresholds (1, 5, 10, 15, …, 35) were considered, as shown on the x-axis, whereas for former smokers, quitting durations of 15 and 20 years were considered. Combinations of quitting duration and target gender among ever-smokers are color coded, as indicated in the legend on the right.

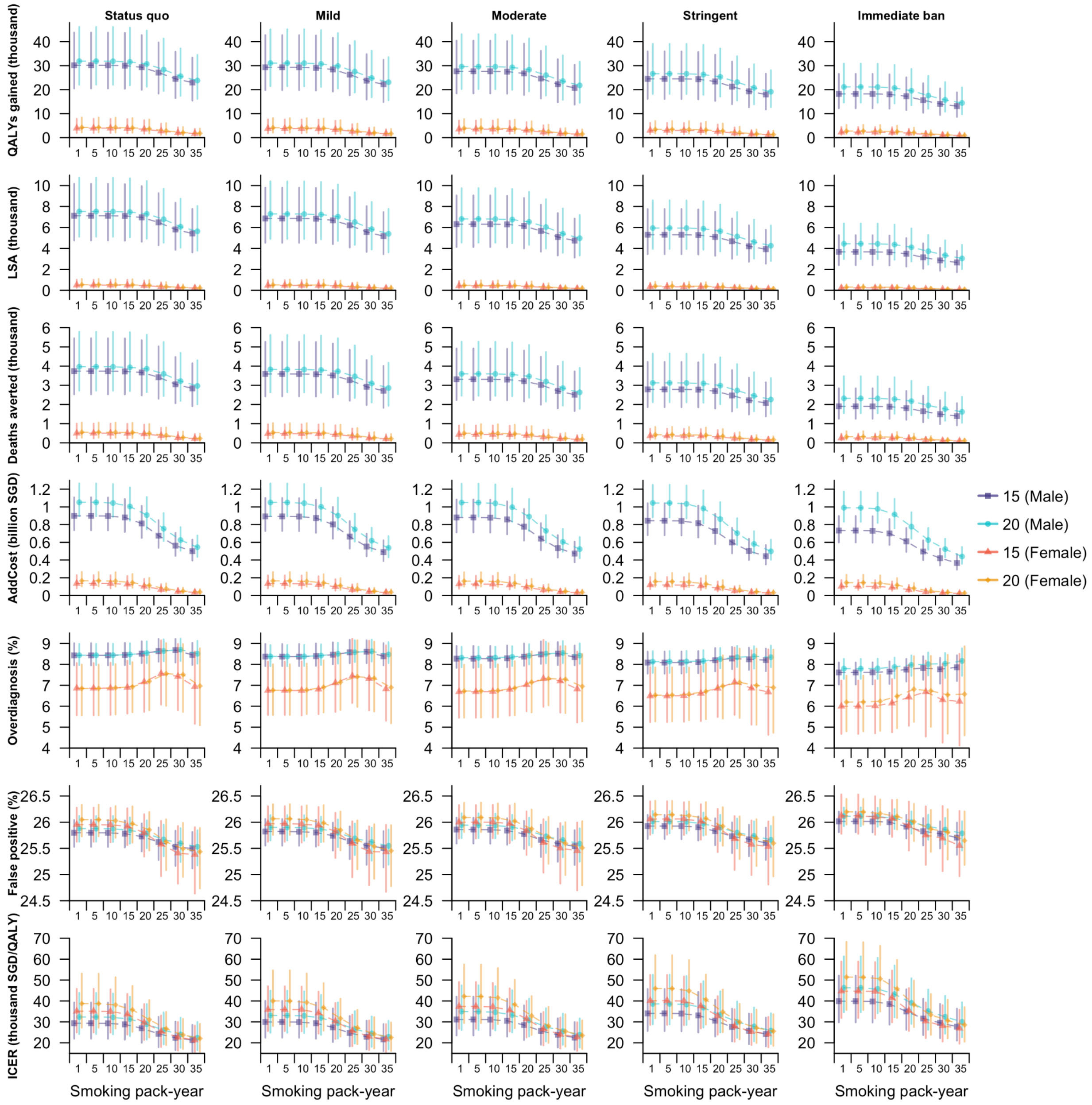

**Figure S14. Projected health and economic impacts of TRS when varying screening eligibility criteria for ever-smokers.** Outcomes of the screening strategies targeting all ever-smokers aged 60–80 years with biennial screening frequency were evaluated over 2025–2050 using seven metrics (one per row), including QALY gains, late-stage cancers averted (LSA), deaths averted, additional costs (AddCost), overdiagnosis rate, false positive rate, and ICER. All these metrics were calculated related to the no-additional-screening scenario. For each metric, dots represent the median across simulations, and vertical lines indicate the corresponding 95% uncertainty intervals (UI). Columns correspond to the five tobacco control scenarios. For current smokers, eight pack-year thresholds (1, 5, 10, 15, …, 35) were considered, as shown on the x-axis, whereas for former smokers, quitting durations of 15 and 20 years were considered. Combinations of quitting duration and target gender among ever-smokers are color coded, as indicated in the legend on the right.

**Sensitivity analysis: Screening individuals with a family history**

We additionally assessed the health and economic impacts of extending screening to individuals with a family history (defined as having at least one first-degree relative diagnosed with lung cancer), who comprised approximately 3% of the population. For ever-smokers (defined as current smokers with at least 20 pack-years of smoking exposure and former smokers who had quit fewer than 15 years earlier), we assumed either annual screening for all aged 50–80 years or biennial screening for all aged 60–80 years, in accordance with MLSOD10 and TRS identified in the main analysis, respectively. For individuals with a family history who did not meet these eligibility criteria, we applied all screening strategies examined in the main analysis, but with a screening frequency no higher than that for the eligible ever-smokers (i.e., annual screening was not considered when applying TRS for ever-smokers). This group comprised never-smokers, current smokers with fewer than 20 pack-years, and former-smokers who had quit more than 15 years earlier. Stage-specific screening sensitivities also varied according to smoking status, with values informed by literature and empirical evidence (Table S10).[7,8]

**Table S10. Screening sensitivity by smoking status.**

| Stage | Ever smokers (Main analysis) | Never smokers |
|---|---|---|
| I | 0.40 | 0.53 |
| II | 0.44 | 0.56 |
| III | 0.75 | 0.76 |
| IV | 0.98 | 0.98 |

We estimated the incremental costs and QALYs associated with extending screening and treating individuals with a family history of lung cancer, in addition to ever-smokers eligible for MLSOD10 or TRS in each tobacco control scenario, and derived ICERs for screening this population accordingly. The additional QALY gains and costs from this expansion in the screening population was marginal (<5%) relative to screening eligible ever-smokers (Figure S15–S16).

In both cases, screening females with a family history generated substantially greater QALY gains than screening males, resulting in much lower ICERs, and screening females aged 55 years and above at a triennial or lower frequency was projected to be cost-effective (i.e., ICER not exceeding SGD 0.12 million per QALY). The QALY gains, additional costs, and ICERs did not vary substantially across tobacco control scenarios (Figure S17–S18).

When MLSOD10 (annual screening of all aged 50–80 years) was adopted for ever-smokers, this strategy was also the most life-saving for the subpopulation with a family history among all strategies with an over-diagnosis rate below 10%. In contrast, the overall optimal strategy for individuals with a family history suggested biennial screening of all males aged 50–80 years and females aged 55–80 years in most tobacco control scenarios. The only exception was the immediate ban scenario, in which the recommended screening age range for males was narrowed to 50–75 years (Figure S19).

When TRS (biennial screening of all aged 60–80 years) was adopted for ever-smokers, the most life-saving strategy with an overdiagnosis rate below 10% was biennial screening of all males with a family history aged 50–80 years and triennial screening of all females with a family history aged 50–85 years. In contrast, the overall optimal strategy was biennial screening of all eligible males and females aged 55–80 years across all scenarios except the immediate ban scenario, where the eligible age range for males was expanded to 55–75 years (Figure S20).

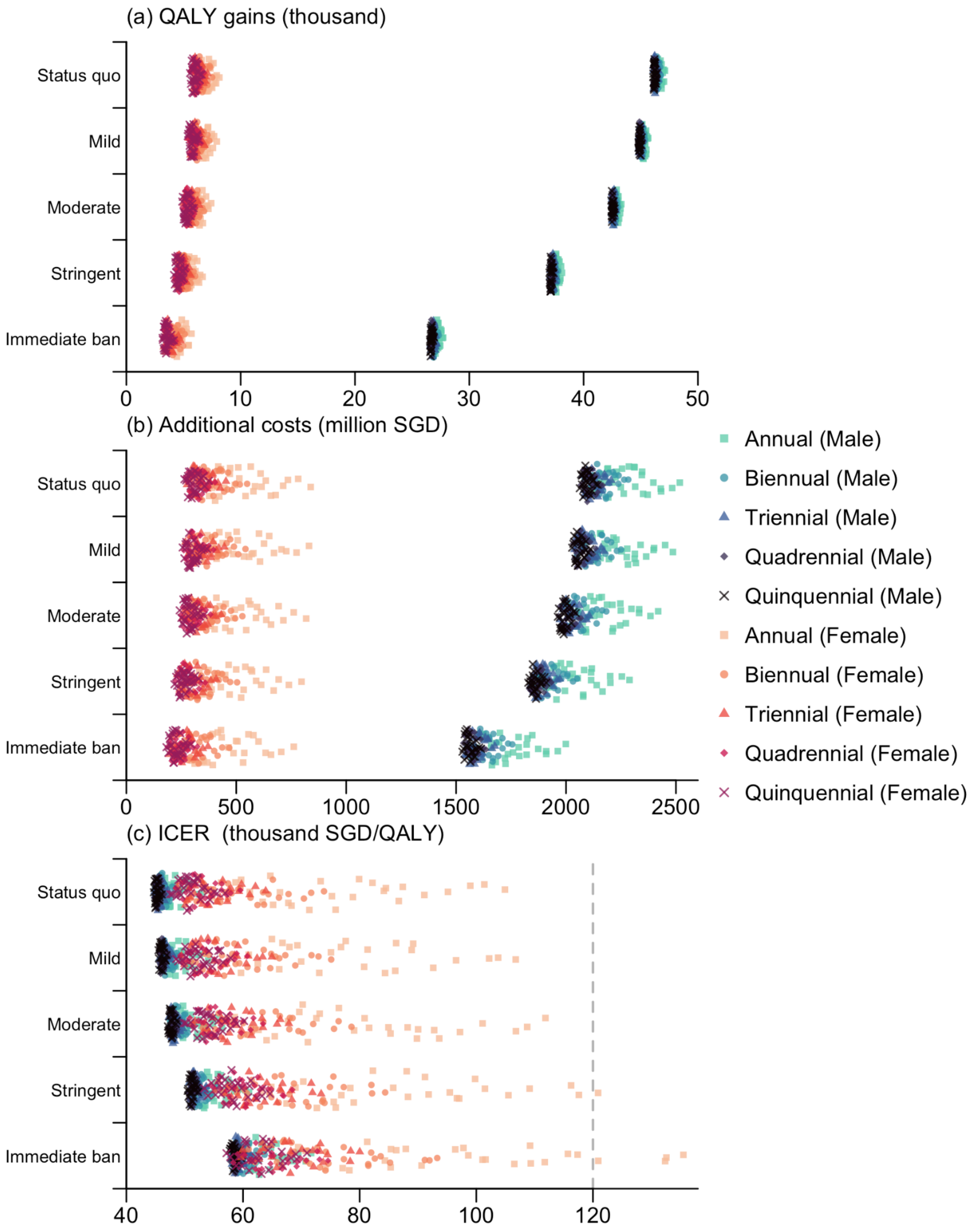

**Figure S15. QALYs gained, costs incurred, and ICERs associated with screening both eligible ever-smokers and individuals with a family history, when eligible ever-smokers were screened following MLSOD10 (annual for all aged 50–80 years).** Differences in these health and economic outcomes were calculated by comparing against the no-additional-screening scenario within each microsimulation across five tobacco control settings.

Strategies are characterised by target populations, screening frequencies, and uptake levels, with combinations of frequency and target gender represented by different colors and point styles (as shown in the legend on the right). The dashed line in subfigure (c) represents the willingness-to-pay threshold of 0.12 million SGD per QALY.

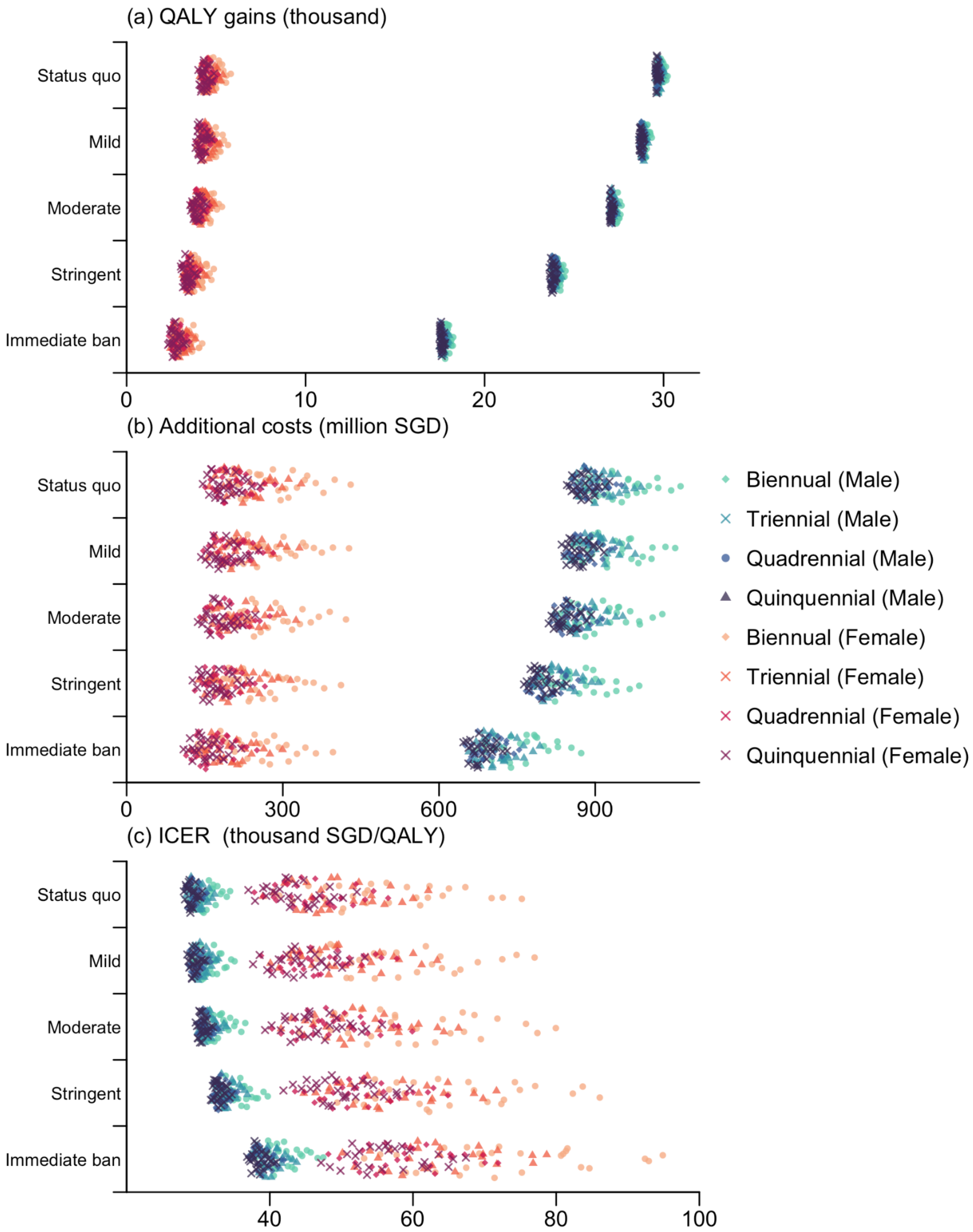

**Figure S16. QALYs gained, costs incurred, and ICERs associated with screening both eligible ever-smokers and individuals with a family history, when eligible ever-smokers were screened following TRS (biennial for all aged 60–80 years).** Differences in these health and economic outcomes were calculated by comparing against the no-additional-screening scenario within each microsimulation across five tobacco control settings. Strategies are characterised by target populations, screening frequencies, and uptake levels, with combinations of frequency and target gender represented by different colors and point styles (as shown in the legend on the right). The dashed line in subfigure (c) represents the willingness-to-pay threshold of 0.12 million SGD per QALY.

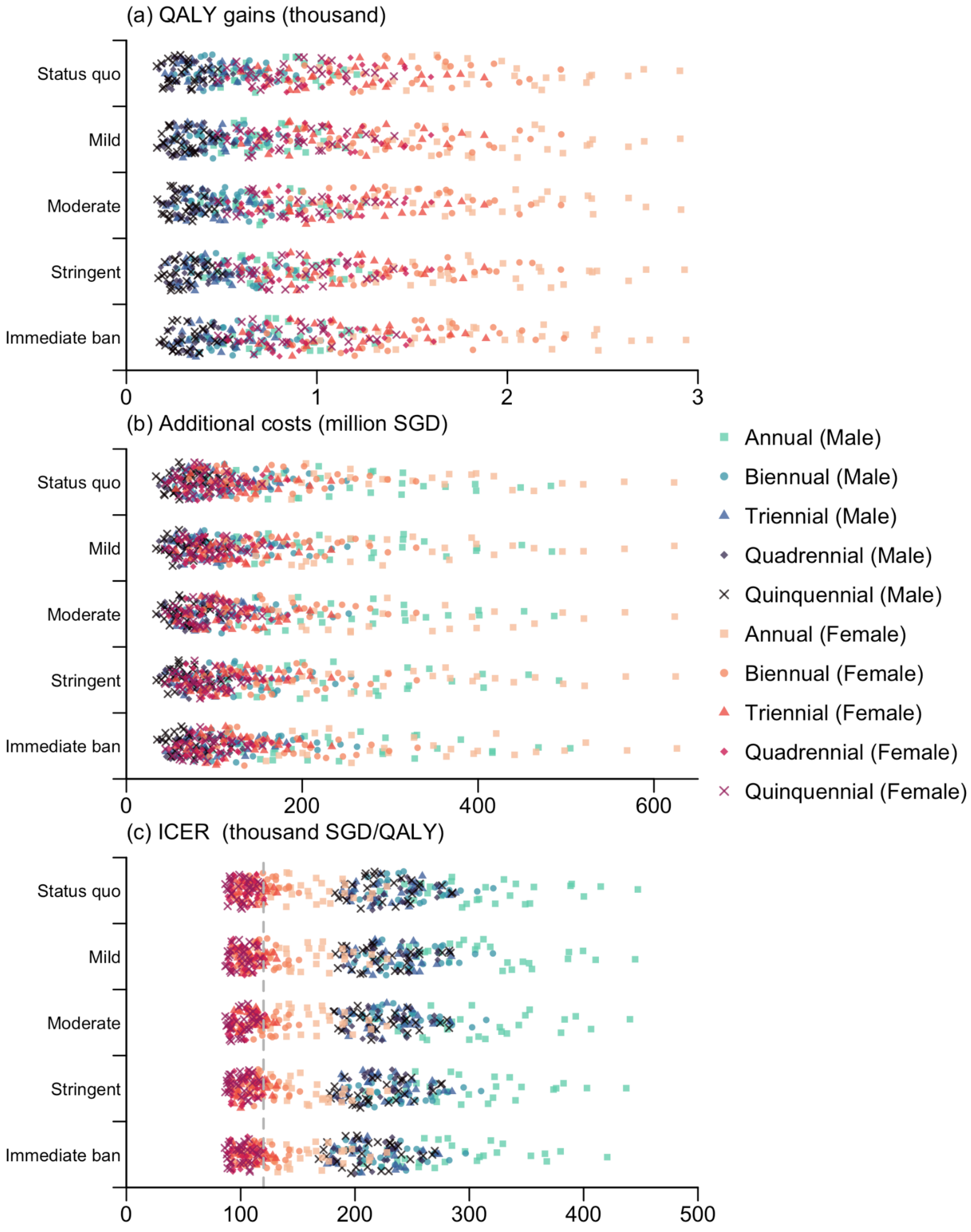

**Figure S17. QALYs gained, costs incurred, and ICERs associated with additionally screening individuals with a family history when eligible ever-smokers were screened following MLSOD10 (annual for all aged 50–80 years).** Differences in these health and economic outcomes were calculated by comparing strategies that additionally screened individuals with a family history against TRS that exclusively screened ever-smokers within each microsimulation across five tobacco control scenarios. Strategies are characterised by target populations, screening frequencies, and uptake levels, with combinations of frequency and target gender represented by different colors and point styles (as shown in the legend on the right). The dashed line in subfigure (c) represents the willingness-to-pay threshold of 0.12 million SGD per QALY.

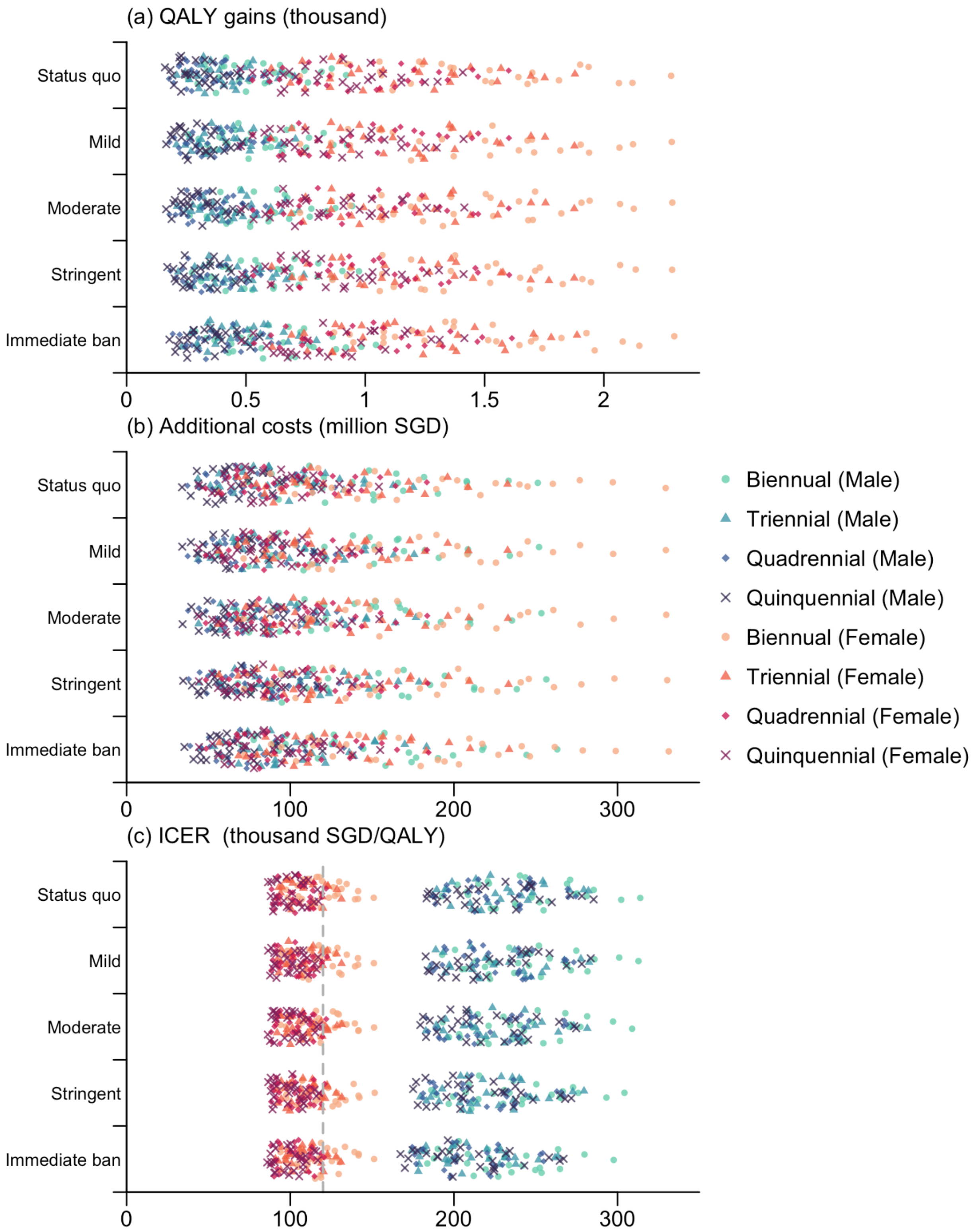

**Figure S18. QALYs gained, costs incurred, and ICERs associated with additionally screening individuals with a family history when eligible ever-smokers were screened following TRS (biennial for all aged 60–80 years).** Differences in these health and economic outcomes were calculated by comparing strategies that additionally screened individuals with a family history against TRS that exclusively screened ever-smokers within each microsimulation across five tobacco control scenarios. Strategies are characterised by target populations, screening frequencies, and uptake levels, with combinations of frequency and target gender represented by different colors and point styles (as shown in the legend on the right). The dashed line in subfigure (c) represents the willingness-to-pay threshold of 0.12 million SGD per QALY.

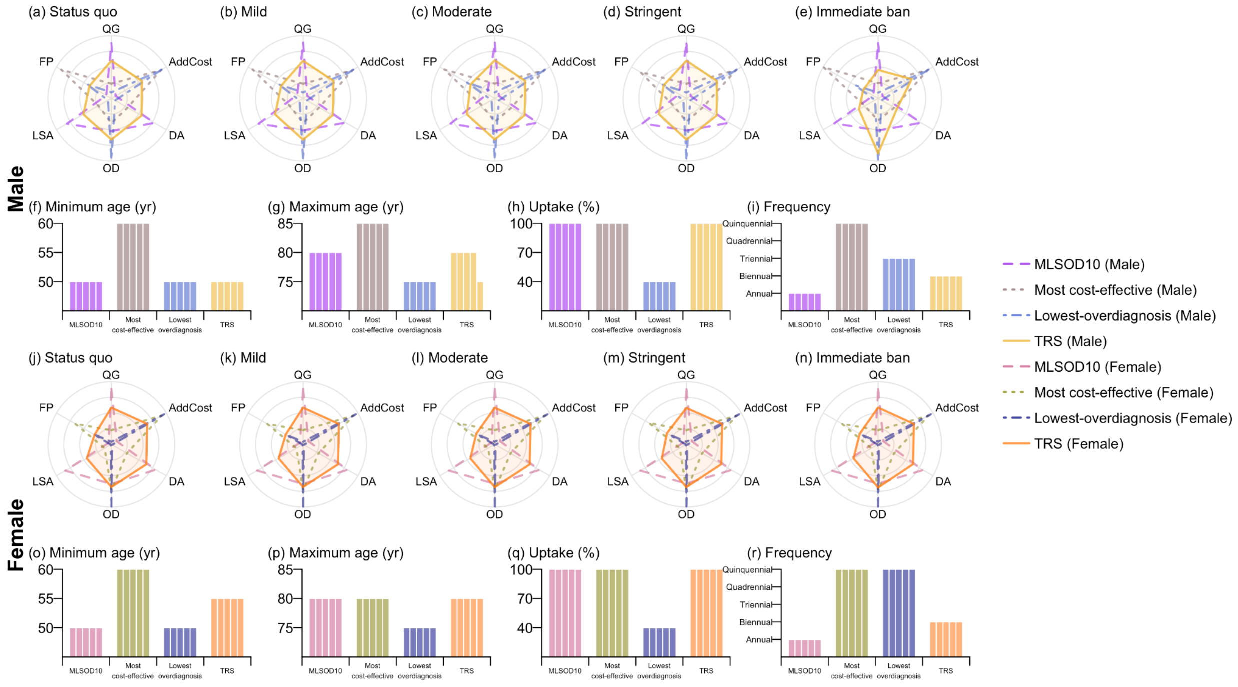

**Figure S19. Recommended screening strategies for screening individuals with a family history under four decision criteria when eligible ever-smokers were screened following MLSOD10 (annual for all aged 50–80 years).** The four selected strategies comprise the most life-saving with 10% overdiagnosis rate (MLSOD10), the most cost-effective, the lowest-overdiagnosis, and the overall optimal strategy (TRS). Subfigures (a)–(e) (Row 1, for males) and (j)–(n) (Row 3, for females) present the scores for six metrics evaluated over year 2025–2050: QALY gains (QG), late-stage cancer averted (LSA), deaths averted (DA), and additional costs (AddCost) relative to the scenario without additional screening, as well as overdiagnosis rate (OD) and false positive rate (FP). All the metrics represent medians across simulations and are rescaled to a 0–1 range. Subfigures (f)–(i) (Row 2, for male ever-smokers) and (o)–(r) (Row 4, for female ever-smokers) show the characteristics of the selected strategies, including minimum age, maximum age, screening uptake rate, and screening frequency (rescaled to an annual rate). The five adjacent columns within each decision criteria correspond to the five tobacco control scenarios ranging from status quo to immediate ban. Strategies selected under different criteria for males and females are distinguished by colour and line style, as indicated in the legend on the right.

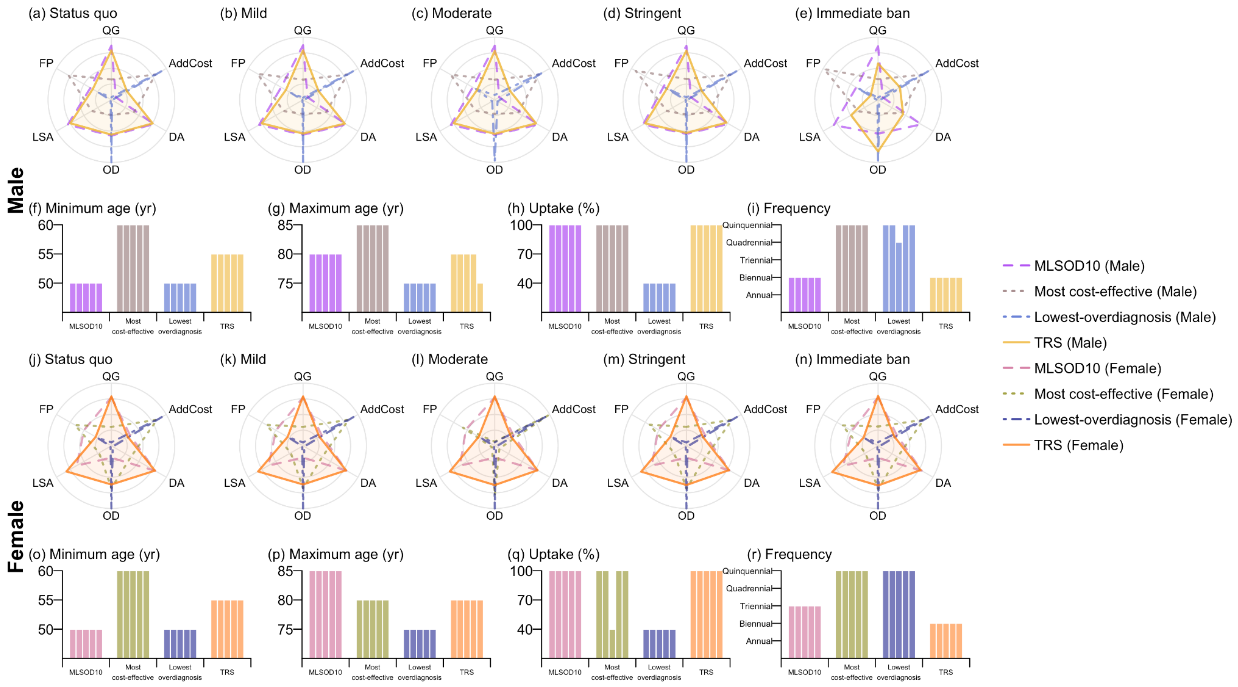

**Figure S20. Recommended screening strategies for screening individuals with a family history under four decision criteria when eligible ever-smokers were screened following TRS (biennial for all aged 60–80 years).** The four selected strategies comprise the most life-saving with 10% overdiagnosis rate (MLSOD10), the most cost-effective, the lowest-overdiagnosis, and the overall optimal strategy (TRS). Subfigures (a)–(e) (Row 1, for males) and (j)–(n) (Row 3, for females) present the scores for six metrics evaluated over year 2025–2050: QALY gains (QG), late-stage cancer averted (LSA), deaths averted (DA), and additional costs (AddCost) relative to the scenario without additional screening, as well as overdiagnosis rate (OD) and false positive rate (FP). All the metrics represent medians across simulations and are rescaled to a 0–1 range. Subfigures (f)–(i) (Row 2, for male ever-smokers) and (o)–(r) (Row 4, for female ever-smokers) show the characteristics of the selected strategies, including minimum age, maximum age, screening uptake rate, and screening frequency (rescaled to an annual rate). The five adjacent columns within each decision criteria correspond to the five tobacco control scenarios ranging from status quo to immediate ban. Strategies selected under different criteria for males and females are distinguished by colour and line style, as indicated in the legend on the right.
